# Identification of disease-specific TCRs maintaining pathogenic T helper cell responses in postinfectious Lyme Arthritis

**DOI:** 10.1101/2024.01.18.24301314

**Authors:** Johannes Dirks, Jonas Fischer, Julia Klaussner, Christine Hofmann, Annette Holl-Wieden, Viktoria Buck, Christian Klemann, Hermann Girschick, Ignazio Caruana, Florian Erhard, Henner Morbach

## Abstract

**Background:** Antibiotic-Refractory Lyme Arthritis(ARLA) involves a complex interplay of T cell responses targeting *Borrelia burgdorferi* antigens succeeding towards autoantigens by epitope spreading. However, the precise molecular mechanisms driving the pathogenic T cell response in ARLA remain unclear. Our aim was to elucidate the molecular program of disease-specific Th cells.

**Methods:** Using flow cytometry, high-throughput T cell receptor (TCR) sequencing and scRNA-seq of CD4^+^ Th cells isolated from the joints of European ARLA patients, we aimed at inferring antigen specificity through unbiased analysis of TCR repertoire patterns, identifying surrogate markers for disease-specific TCRs and connecting TCR specificity to transcriptional patterns.

**Results:** PD-1^hi^HLA-DR^+^CD4^+^ effector T cells were clonally expanded within the inflamed joints and persisted throughout disease course. Among these cells, we identified a distinct TCRβ motive restricted to HLA-DRB1*11 or *13 alleles. These alleles, being underrepresented in North American ARLA patients, were unexpectedly prevalent in our European cohort. The identified TCRβ motive served as surrogate marker for a convergent TCR response specific to ARLA, distinguishing it from other rheumatic diseases. In the scRNA-seq dataset, the TCRβ motive particularly mapped to peripheral T helper (T_PH_) cells displaying signs of sustained proliferation, continuous TCR signaling, and expressing CXCL13 and IFN-γ.

**Conclusion:** By inferring disease-specific TCRs from synovial T cells we identified a convergent TCR response in the joints of ARLA patients that continuously fueled the expansion of T_PH_ cells expressing a pathogenic cytokine effector program. The identified TCRs will aid in uncovering the major antigen targets of the maladaptive immune response.

**Funding:** Supported by the German Research Foundation (DFG) MO 2160/4-1; the Federal Ministry of Education and Research (BMBF; Advanced Clinician Scientist-Program INTERACT; 01EO2108) embedded in the Interdisciplinary Center for Clinical Research (IZKF) of the University Hospital Würzburg; the German Center for Infection Research (DZIF; Clinical Leave Program; TI07.001_007) and the Interdisciplinary Center for Clinical Research (IZKF) Würzburg (Clinician Scientist Program, Z-2/CSP-30).

## INTRODUCTION

Lyme disease, caused by the tick-transmitted spirochete *Borrelia (B.) burgdorferi sensu lato*, is the most prevalent vector-borne illness in North America and Europe, posing a significant public health concern (1, 2). In North America, *B. burgdorferi sensu stricto* (s.s.) is the main species, while in Europe, additional species such as *B. afzelii* and *B. garinii* contribute to disease burden (3). Among the late manifestations, Lyme Arthritis (LA) is the most commonly reported (4). While the majority of cases resolve after antibiotic treatment, a small subgroup of individuals experiences persistent joint inflammation despite adequate antibiotic treatment named postinfectious or antibiotic-refractory Lyme arthritis (ARLA) (5–8).

ARLA is characterized by an exaggerated proinflammatory immune response that persists after the initial infection resulting in synovitis and synovial hyperplasia. *Borrelia* outer-surface protein A (OspA) has been identified as one of the major targets eliciting this dysregulated immune responses (9–12). The risk of developing ARLA has been linked to specific HLA-DRB1 alleles that efficiently present OspA-derived peptides to CD4^+^ T cells (11, 13). In a North American study, HLA-DRB1 alleles *01:01, *04:01, and *15:01 were associated with a higher risk, while *08:01, *11:01, *11:04, and *13:02 were associated with a potential protective capacity (13). In individuals at risk, OspA appears to trigger an excessive immune response creating a highly inflammatory milieu that subsequently promotes epitope spreading of the immune response towards autoantigens (7). This autoimmune response seems to be mediated by T-bet and/or RORγt expressing Th cells and target extracellular matrix proteins as well as vascular autoantigens (14–18).

While this may point towards Th1 and/or Th17 cells as drivers of arthritis, the detailed function and molecular program of pathogenic Th cells in ARLA patients remain unclear. Moreover, the concept of an OspA-mediated dysregulated immune response might not directly extend to regions outside North America where the composition of *Borrelia* species causing LA differs. This discrepancy is evident since immune responses against OspA are noticeable in many North American patients, especially those experiencing ARLA, but they are seldom observed in European patients with LA (19, 20).

Hence, our objective was to ascertain the molecular program of disease-specific Th cells in patients with ARLA, link TCR specificity to cellular function and track clonal evolution of these cells at the site of inflammation throughout disease progression. Acknowledging the uncertainty of potential target antigens among patients outside North America, we implemented an unbiased approach focusing on the hosts’ immune response. The objective was to infer putative TCR specificity through the clustering of TCRs displaying biological similarities. For this, we employed high-throughput TCR sequencing of activated CD4^+^ Th cells from the joints of patients in conjunction with scRNA-seq.

Using this approach, we identified disease-specific TCRs in inflamed joints of Central European ARLA patients, mainly restricted to HLA-DRB1*11 or HLA-DRB1*13 alleles, which were unexpectedly prevalent in our cohort despite their underrepresentation in the North American cohort. These ARLA-specific T cells exhibited sustained proliferation, likely due to locally induced TCR signaling, and expressed a pathogenic effector program resembling peripheral Th (T_PH_) cells, marked by CXCL13 and IFN-γ expression.

## RESULTS

### PD-1^hi^HLA-DR^+^CD4^+^ effector cells are expanded in the joints of ARLA patients throughout disease course

We studied a cohort of 13 pediatric patients with ARLA (mean ± SD age of onset 13.6 ± 2.2 years; 38.5% female) residing throughout Germany, from whom synovial fluid (SF) was preserved following therapeutic joint injections after they had completed antibiotic therapy. Despite receiving prior adequate antibiotic treatment, all patients presented with chronic arthritis. All patients exhibited a broad serological response against multiple *Borrelia* antigens; however, the majority did not show detectable responses to OspA (Supplemental Table 1). Some patients carried the well-known ARLA HLA-DRB1 risk alleles *01:01, *04:01 and *15:01 including the only patient with a detectable antibody response against OspA and being homozygous for the *15:01 allele. However, five out of the 13 patients (38.5%) carried at least one HLA-DRB1*11 allele, which was underrepresented in the North American ARLA cohort (Supplemental Table 1) (8). In our cohort, the cumulative frequency of the group of known HLA-DRB1 “risk alleles” tended to be lower in comparison to the reported North American cohort (23.1 vs. 38.1%, p=0.18), whereas the frequency of "protective alleles” was significantly higher (42.5 vs. 11.9%, p < 0.01; Supplemental Table 2). Thus, despite the similarity in HLA-DRB1 allele distribution among geographically and ethnically matched control cohorts (Supplemental Table 2), these allele groups exhibited contrasting patterns between the two patient cohorts.

Effector T cells present an instructive cell subset for investigating ongoing immune responses, as they arise from the division of antigen-specific T cells upon TCR activation and are enriched with antigen-specific clones. Recent evidence suggests that disease-specific effector Th cells can be distinguished based on the expression of distinct activation markers (21). To analyze the distribution of recently antigen-activated effector CD4^+^ T cells in the inflamed joints of patients with ARLA, we utilized flow cytometry and employed high expression levels of PD-1 and HLA-DR as surrogate markers. A significant proportion of PD-1^hi^HLA-DR^+^CD4^+^ T cells was observed in the SF of ARLA patients, and these cells exhibited enrichment in CD45RO^+^CCR7^-^ effector memory phenotype (Figure 1A, B). PD1^hi^HLA-DR^+^CD4^+^ T cells were observed in comparable frequencies in both SF and matched synovial tissue samples, with a substantial clonal overlap evident between these compartments (Supplemental Figure 1A-C). Immunohistochemistry analysis revealed synovial hyperplasia and synovial infiltration of PD-1 expressing CD4^+^ T cells that formed aggregates with CD20^+^ B cells (Figure 1C, Supplemental Figure 1D). Analyzing the clonal diversity by sequencing the TCRβ repertoire within bulk sorted SF PD-1^hi^HLA-DR^+^ and PD-1l^o/-^HLA-DR^-^ CD4^+^ T cells revealed a more restricted repertoire in the PD-1^hi^HLA-DR^+^ subset as an indicator of oligoclonal expansion (Figure 1D). Additionally, the PD-1^hi^HLA-DR^+^CD4 T cell subset demonstrated a significant enrichment in cells expressing Ki-67 as sign of ongoing proliferation (Figure 1E, F, p<0.01).

**Figure 1.**
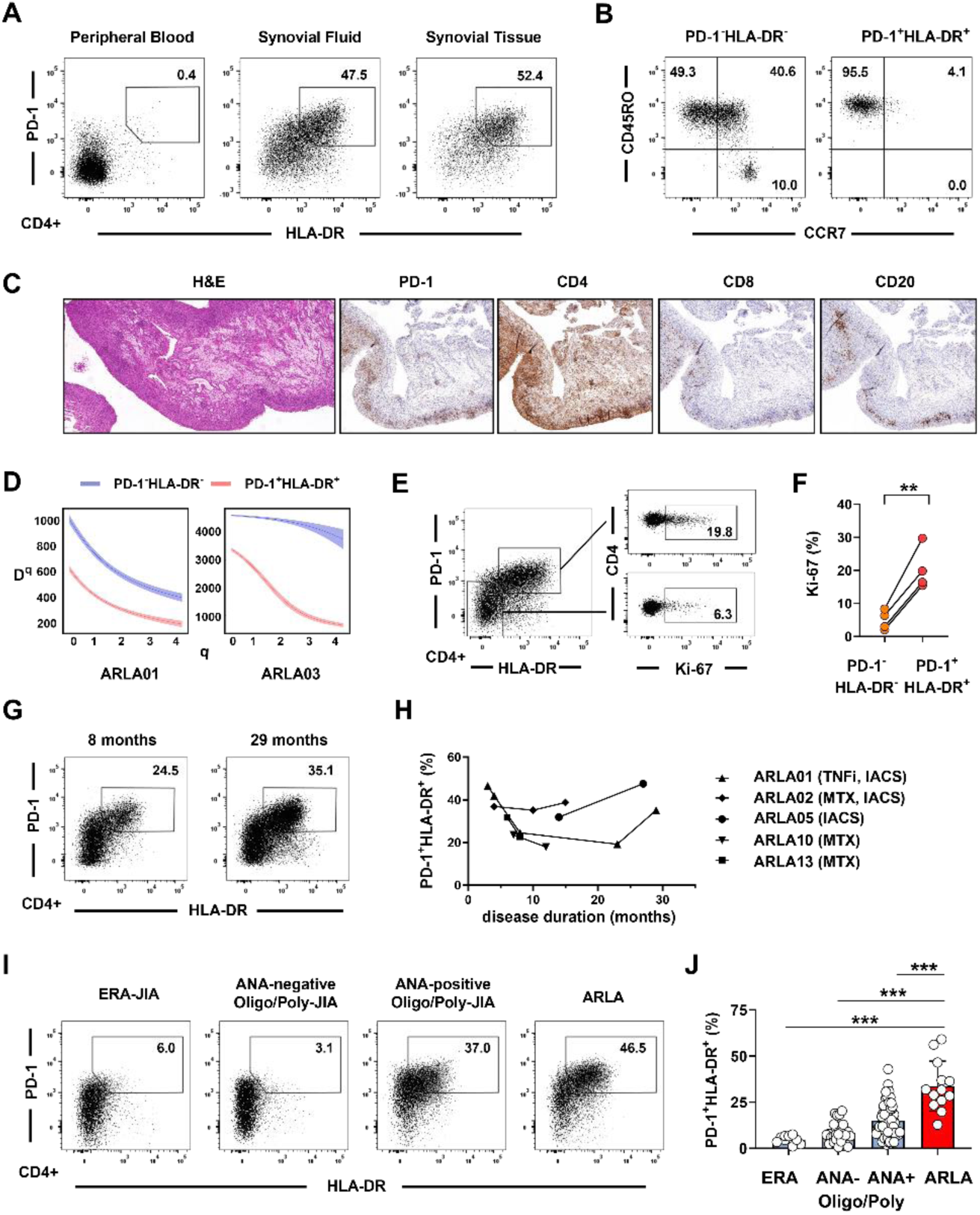
Oligoclonal expansion of PD-1^hi^HLA-DR^+^CD4^+^ effector T cells in the joints of ARLA patients throughout disease course. **(A)** Representative dot plots showing PD-1 and HLA-DR expression on matched peripheral blood (PB), synovial fluid (SF) and synovial tissue (ST) CD4^+^ T cells from patient ARLA05. **(B)** Dot plots presenting CCR7 and CD45RO expression on SF PD-1^hi^HLA-DR^+^ and PD-1^lo/-^HLA-DR^-^ CD4^+^ T cells. **(C)** Representative histological and immunohistological analysis of ST sections from patient ARLA01. (**D)** Clonal diversity analysis of the TCR Vβ repertoire within both T cell populations from 2 ARLA patients. The generalized diversity index (Hill) was computed across a diversity order (q) range using uniform resampling to correct for sequencing depth. The diversity index (qD) is depicted as a smooth curve. **(E)** Dot plot and histogram demonstrating K-i67 expression in PD-1^hi^HLA-DR^+^ and PD-1^lo/-^HLA-DR^-^ SF CD4^+^ T cells. **(F)** Compiled data from four ARLA patients, indicating Ki-67^+^ cell frequencies within the specified populations. Bars represent mean frequency ± standard deviation. Paired two-tailed Student’s t-test, **: p<0.01. **(G)** Dot plots showing PD-1 and HLA-DR expression on SF CD4^+^ T cells from patient ARLA02 at different time points. **(H)** Distribution of SF PD-1^hi^HLA-DR^+^CD4^+^ T cell frequencies in five ARLA patients observed throughout the disease course; intraarticular corticosteroids (IACS), TNF-α inhibitor (TNFi), methotrexate (MTX). **(I)** Dot plots demonstrating PD-1 and HLA-DR expression on SF CD4+ T cells from patients with juvenile idiopathic arthritis (JIA) and ARLA. **(J)** Distribution of SF PD-1^hi^HLA-DR^+^CD4^+^ T cell frequencies stratified based on disease subgroup and/or antinuclear antibody (ANA) status of JIA and ARLA patients. ERA, enthesitis-related arthritis. Bars indicate mean frequency ± standard deviation; Dunnett’s multiple comparisons test from ordinary one-way ANOVA, ***: p<0.001.

We were able to track the expansion of PD-1^hi^HLA-DR^+^CD4^+^ T cells during the disease course in five ARLA patients. Remarkably, a high frequency of PD-1^hi^HLA-DR^+^CD4^+^ T cells persisted in their joints for up to 2.5 years after onset of arthritis, correlating with ongoing synovitis despite prior antibiotic treatment and concurrent anti-inflammatory medication (Figure 1G, H). The frequencies of PD-1^hi^HLA-DR^+^CD4^+^ T cells in the SF of patients with ARLA were found to be significantly higher compared to age-matched patients with various subtypes of juvenile idiopathic arthritis (JIA), which served as disease controls (Figure 1I, J). Notably, the frequency of PD-1^hi^HLA-DR^+^CD4^+^ T cells in ARLA SF even exceeded that observed in patients with antinuclear antibody positive JIA, a condition in which a local autoimmune response is suggested to drive T effector cell expansion (22).

These findings underscore the potential significance of PD-1^hi^HLA-DR^+^CD4^+^ T cells as locally induced effector cells and highlight their promising utility in dissecting the pathogenic T cell response within the inflamed joints of ARLA patients.

### The TCR repertoire of PD-1^hi^HLA-DR^+^ CD4^+^ T cells in the joints of ARLA patients displays signs of an ongoing and convergent T cell response

We speculated whether the sustained expansion of PD-1^hi^HLA-DR^+^CD4^+^ T cells in ARLA patients throughout the disease course might be driven by persistent recognition of disease specific antigens. To investigate this, we first analyzed the TCR repertoire of these cells for indicative markers. Considering HLA restriction and the unexpectedly high frequency of the HLA-DRB1*11 allele in our ARLA cohort, we initially focused our analysis on patients who carried at least one HLA-DRB1*11 allele (Supplemental Table 1 and 3). To comprehensively assess the TCR repertoire on a larger scale, we sorted SF PD-1^hi^HLA-DR^+^CD4^+^ T cells from five patients and analyzed their TCRβ repertoire by bulk sequencing. Each sample yielded between 529 and 4774 distinct clones, with minimal clonal overlap observed among the analyzed individuals (0, 0 or 5 shared clonotypes between 5, 4 or 3 individuals, respectively). As this approach failed to reveal significant numbers of shared clonotypes indicative of a disease-specific TCR response, we next utilized GLIPH2 (Grouping of Lymphocyte Interactions by Paratope Hotspots, version 2) (23). This algorithm clusters TCRs based on shared sequence similarities rather than identities, predicting them to bind the same MHC-restricted peptide antigen. By applying this method to the combined set of bulk TCRβ sequences, we identified 593 different specificity groups overlapping between at least two individuals (Figure 2A). Performing a network visualization of those sequences and their specificity groups, we identified a cluster of significantly enriched TCRs that were closely linked by similar specificity groups (highlighted as ’Specificity Cluster’ in Figure 2B). This cluster comprised 3.0 to 6.7% of the total clonal space and was enriched with specificity groups that shared similar local motives located in the part of the CDR3β not coded for by germline *TRBV* and *TRBJ* segments (therefore n-/p-nucleotides, Table 1).

**Figure 2.**
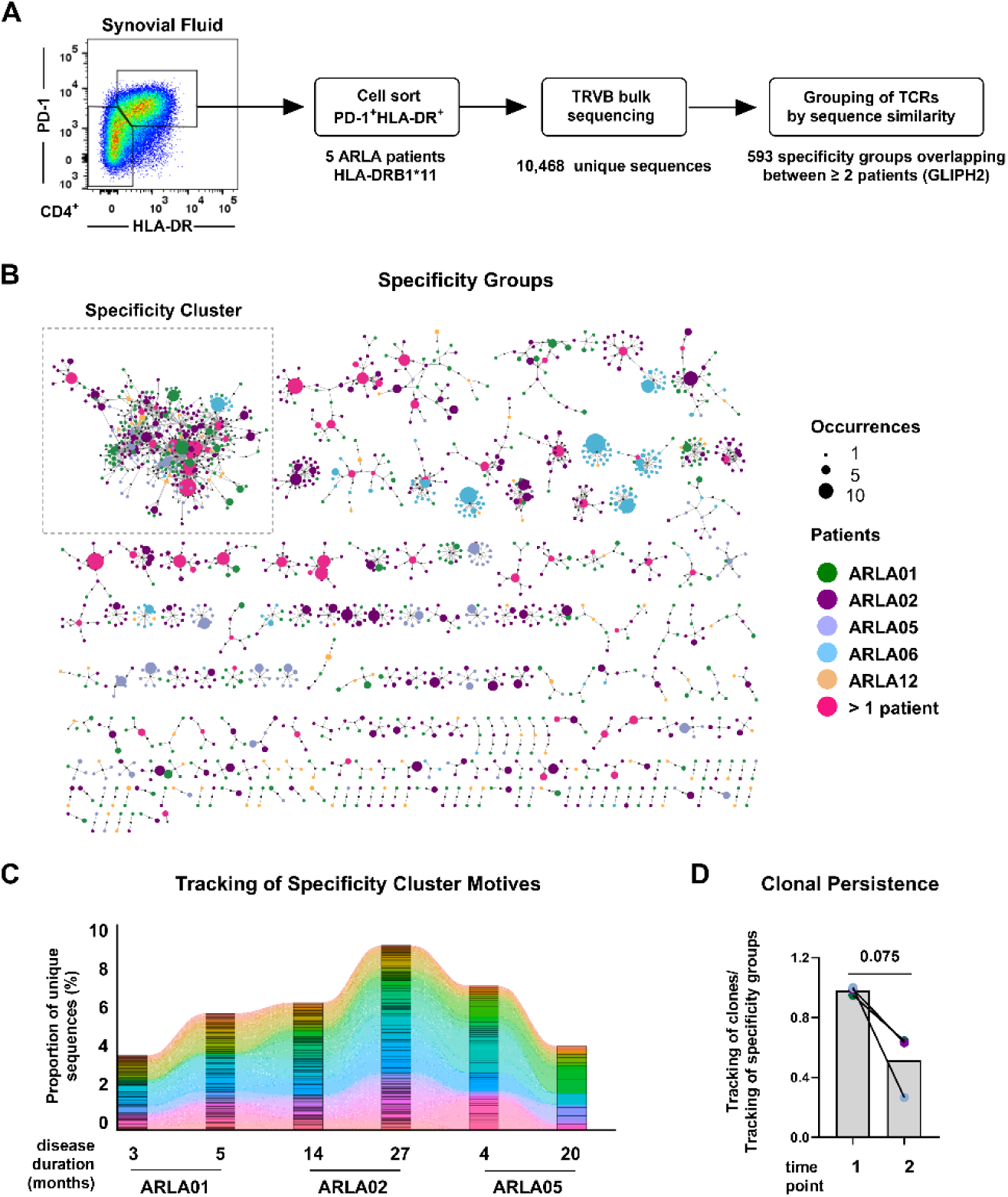
– Convergent and ongoing T cell responses in the joints of ARLA patients. **(A)** Schematic work-flow illustrating the analysis process for identification of TCR similarities and clustering of TCRs into groups based on their probable specificity. **(B)** Network representation displaying TCR specificity groups enriched by GLIPH2 in SF PD-1^hi^HLA-DR^+^CD4^+^ T cells from five ARLA patients with at least one HLA-DRB1*11 allele. Only specificity groups containing sequences from multiple patients are shown. Motives are represented by small black circles, and corresponding CDR3β sequences as colored circles; colors correspond to the sourcing individual sizes indicate the absolute abundancies of unique CDR3 amino acid (aa) sequences in all patients. **(C)** Tracking of occupied repertoire space within SF PD-1^hi^HLA-DR^+^CD4^+^ T cells using sequences containing CDR3 aa motives from the ‘specificity cluster’ at various time points in three patients. Each color corresponds to an unique CDR3 aa sequence. **(D)** Ratio comparison of the occupied repertoire space by sequences containing CDR3 aa motives from the ‘specificity cluster’, as defined in (B), against the occupied repertoire space by CDR3 aa sequences in the ‘specificity cluster’ at time point 1 (as depicted in Supplemental Figure 2A); paired two-tailed Student’s t-test.

**Table 1.** CDR3β sequence and VJ gene usage contained within the ‘specificity cluster’s.

| Pattern | Unique CDR3s | Most frequent unique CDR3s |  |  | Number of unique sequences |  |  |  |  |
| --- | --- | --- | --- | --- | --- | --- | --- | --- | --- |
|  |  | TRBV | TRBJ | CDR3 aa | ARLA01 | ARLA02 | ARLA05 | ARLA06 | ARLA12 |
| **SLSY** | 61 | TRBV18 | TRBJ2-7 | CASSPIasISYEQYF | 1 | 4 | 5 |  |  |
|  |  | TRBV18 | TRBJ2-7 | CASSPIssISYEQYF | 2 | 7 |  |  |  |
|  |  | TRBV7-2 | TRBJ2-7 | CASSLtasISYEQYF |  | 7 | 2 |  |  |
|  |  | Total sequences / patient in specificity group: |  |  | 38 | 47 | 27 |  | 6 |
| **TLSL** | 38 | TRBV7-2 | TRBJ2-7 | CASSatlsItYEQYF |  | 6 |  |  |  |
|  |  | TRBV7-2 | TRBJ1-1 | CASSLtlslAYvAFF |  | 6 |  |  |  |
|  |  | TRBV7-2 | TRBJ2-7 | CASSStlsItYEQYF |  |  | 4 |  |  |
|  |  | Total sequences / patient in specificity group: |  |  | 11 | 40 | 11 |  |  |
| **SLTL** | 33 | TRBV7-2 | TRBJ1-1 | CASSLtlslAYvAFF |  | 6 |  |  |  |
|  |  | TRBV7-2 | TRBJ2-7 | CASSLtlslSYEQYF | 1 | 3 |  |  |  |
|  |  | TRBV7-2 | TRBJ2-7 | CASSLtlsvtYEQYF |  | 3 |  |  |  |
|  |  | Total sequences / patient in specificity group: |  |  | 8 | 30 | 4 |  | 4 |
| **SLTY** | 29 | TRBV7-2 | TRBJ2-7 | CASSatlsItYEQYF |  | 6 |  |  |  |
|  |  | TRBV7-2 | TRBJ2-7 | CASSSTGsltYEQYF | 5 |  |  |  |  |
|  |  | TRBV7-2 | TRBJ2-7 | CASSStlsItYEQYF |  |  | 4 |  |  |
|  |  | Total sequences / patient in specificity group: |  |  | 22 | 12 | 8 |  |  |
| **SVTY** | 26 | TRBV18 | TRBJ2-7 | CASSPIssvtYEQYF | 9 |  |  |  |  |
|  |  | TRBV18 | TRBJ2-7 | CASSPIasvtYEQYF |  | 4 |  |  |  |
|  |  | TRBV7-2 | TRBJ2-7 | CASSLtlsvtYEQYF |  | 3 |  |  |  |
|  |  | Total sequences / patient in specificity group: |  |  | 19 | 25 |  |  | 1 |
| **LTLS** | 26 | TRBV7-2 | TRBJ1-1 | CASSLtlslAYvAFF |  | 6 |  |  |  |
|  |  | TRBV7-2 | TRBJ2-7 | CASSLtlslSYEQYF | 1 | 3 |  |  |  |
|  |  | TRBV7-2 | TRBJ2-7 | CASSLtlslAYEQYF |  | 3 |  |  |  |
|  |  | Total sequences / patient in specificity group: |  |  | 5 | 27 | 4 |  | 3 |
| **LASV** | 21 | TRBV5-4 | TRBJ2-3 | CASSLasvaDTQYF |  |  |  | 9 |  |
|  |  | TRBV18 | TRBJ2-7 | CASSPLAsvSYEQYF | 4 | 1 |  |  |  |
|  |  | TRBV18 | TRBJ2-7 | CASSPLAsvtYEQYF |  | 3 |  |  |  |
|  |  | Total sequences / patient in specificity group: |  |  | 5 | 11 |  | 19 |  |
| **ASLS** | 20 | TRBV18 | TRBJ2-7 | CASSPIasISYEQYF | 1 | 4 | 5 |  |  |
|  |  | TRBV7-2 | TRBJ2-7 | CASSLtasISYEQYF |  | 2 | 7 |  |  |
|  |  | TRBV18 | TRBJ2-7 | CASSPLAsISYEQYF | 4 |  |  |  | 1 |
|  |  | Total sequences / patient in specificity group: |  |  | 6 | 29 | 9 |  | 3 |

We next sought to investigate the longitudinal kinetics of the ‘specificity cluster’ within the PD-1^hi^HLA-DR^+^ cell subset in SF samples from three ARLA patients collected at various time points during disease course. TCRs carrying ’specificity cluster’ motives remained detectable in subsequent samples at frequencies comparable to those observed during the initial sampling, with clonal persistence even after intervals of up to 16 months between analysis time points (Figure 2C; Supplemental Figure 2A). In addition, nearly half of the TCRs associated with any ’specificity cluster‘ motives in the follow-up samples could not be identified in the initial sample suggesting that new clones with similar TCRs are recruited into the ‘specificity cluster’ over time (Figure 2D and Supplemental Figure 2A). Exploring deeper into the CDR3β sequences corresponding to the most frequent specificity groups in the cluster, we uncovered greater nucleotide diversity compared to the amino acid (aa) level as a characteristic sign of a convergent T cell response (Supplemental Figure 2B, C).

Thus, within SF PD-1^hi^HLA-DR^+^CD4^+^ T cells of ARLA patients exhibiting a distinct HLA-DRB1 background, we identified a persistent cluster of TCR specificity groups that endured throughout the disease course. This cluster was partially replenished over time by different clones with identical TCR motives. This observation aligns with a continually ongoing T cell response in the joints of these ARLA patients, triggered by and converging toward a set of antigens.

### A HLA-DRB1 restricted TCRβ amino acid motive functions as surrogate marker for ARLA specific TCRs

To facilitate the identification of disease-specific TCRs in ARLA, our objective was to uncover surrogate markers capable of comprehensively identifying these TCRs with minimal effort. To achieve this, we conducted a detailed analysis of the fundamental molecular patterns exhibited by the TCRs within the previously defined ‘specificity cluster‘.

Analysis of the TCRβ VJ pairings revealed a significant increase of the TRBV7-2.TRBJ2-7 and TRBV18.TRBJ2-7 combinations in TCRs within the cluster compared to all others (74.8% vs. 1.5%, respectively; p< 0.0001 by Chi square with Yate’s correction; Figure 3A). In addition, TCRβ sequences contributed by patient ARLA06 to the specificity cluster displayed a distinct VJ pairing with predominance of TRBV5-4.J2-3 (Supplemental Figure 3A). The TCRβ sequences from all patients in the cluster almost exclusively used the aa duplet ‘SL’ or ‘SV’ within the hypervariable part of the CDR3 region at IMGT position 111 and 112, which were not encoded by germline template and displayed high variability on the nucleotide level (Table 1, Figure 3B, Supplemental Figure 2B, C and 3B). Additionally, the TCRβ sequences were characterized by usage of ‘GH’ at IMGT position 28 and 29 within the CDR1 region which reflects the use of the above mentioned Vβ segments 7-2, 18 and 5-4 that inherit this motive in their germline configuration (Figure 3B). The simple CDR3β motive (‘SV or SL’) - alone or in combination with the CDR1β motive (‘GH’) - could within the five analyzed patients identify between 55-100% of all TCRs that belonged to the cluster (Figure 3C). Notably, the frequencies of these markers also remained unchanged in SF PD-1^hi^HLA-DR^+^CD4^+^ T cells of ARLA patients during the disease course and were found at similar frequencies in matched synovial tissue (Supplemental Figure 4).

**Figure 3.**
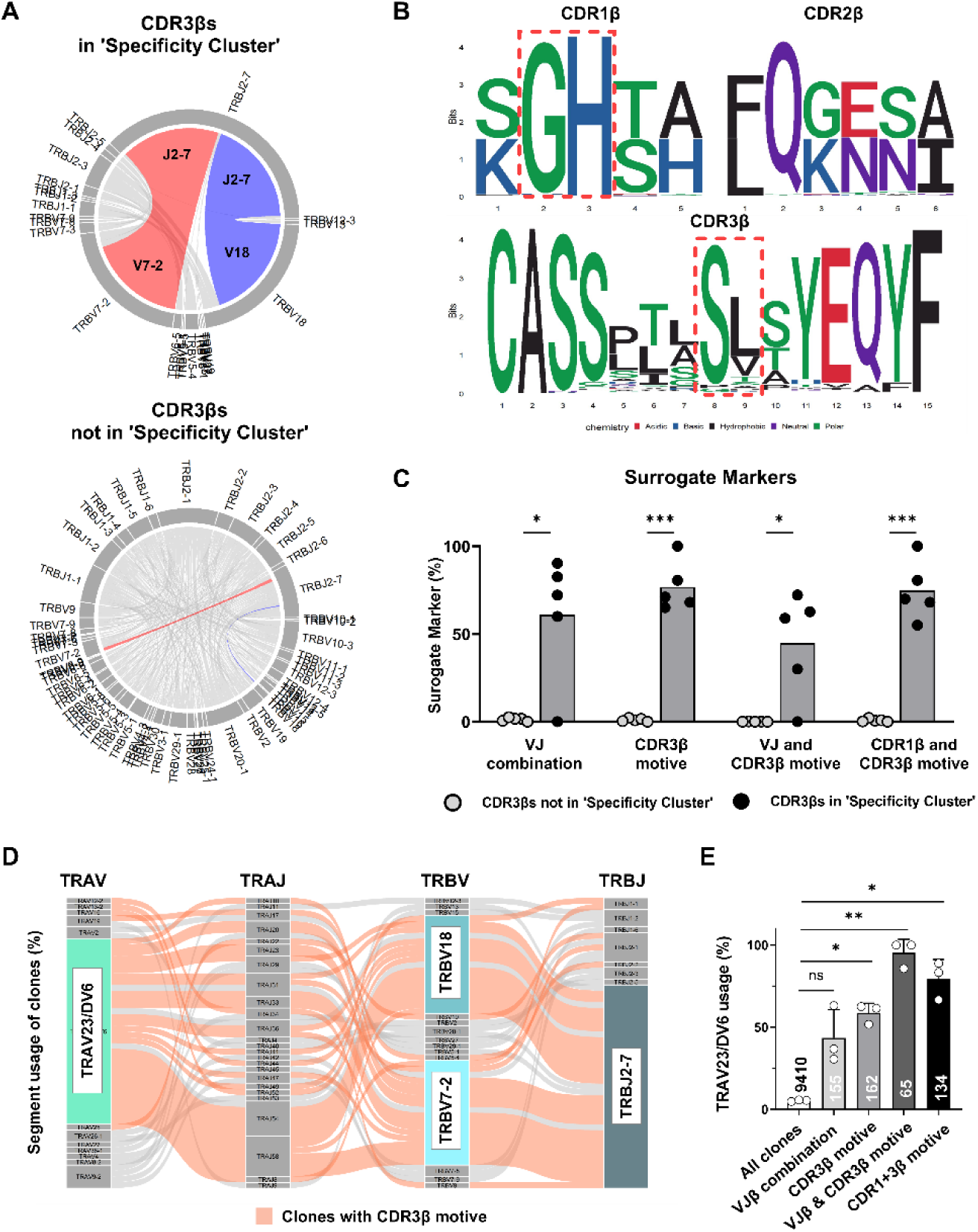
– A combined CDR1β / CDR3β surrogate marker defines a common disease-associated TCRs motives. **(A)** The distribution of TRBV-TRBJ gene segment pairings is depicted in circos plots, showing unique TCRβ chain sequences derived from synovial fluid PD-1^hi^HLA-DR^+^CD4^+^ T cells collected from five ARLA patients. The upper circle delineates sequences belonging to the ’specificity cluster,’ as illustrated in Figure 2A, while the lower circle represents the remaining sequences. The TRBV7-2.TRBJ2-7 and TRBV18.TRBJ2-7 pairing are highlighted in red and blue, respectively. **(B)** Sequence plots depicting the amino acid sequences in CDR1-3β derived from sequences within the ’specificity cluster.’ For the generation of sequence plots, TCR sequences were filtered to include the most abundant length of each CDR. Potential surrogate markers, such as ’GH’ in CDR1β (referred to as ’CDR1β motive’) and ’SL’/’SV’ in CDR3β (referred to as ’CDR3β motive’), are highlighted. **(C)** The frequencies of the indicated surrogate markers are compared between sequences within and outside the ’specificity cluster’; p-values determined by multiple paired t-tests are adjusted for multiple testing by Holm-Šídák method. **(D)** Alluvial plot of TRAV-TRAJ-TRBV-TRBJ combinations (determined by paired TCRα/β-sequencing of CD4^+^ SF T cells from three ARLA patients) in unique clones containing motives from the ‘specificity cluster’ in the CDR3β. Alluvials from clones with the CDR3β motive (‘SL’/’SV’ at IMGT-position 111/112 in CDR3β) are highlighted in coral. **(E)** Frequency of clones with TRAV23/DV6 gene segment usage within all clones or within subsets filtered for the indicated properties of the TCRβ chain. The number of clones in each subset is indicated at each bar. Circles represent individual patients, error bars indicate standard deviation. Significances were calculated by one-way ANOVA and multiple comparisons (to ‘all clones’) corrected with Dunnetts formula; *: p<0.05, **: p<0.01.

We then aimed at including the TCRα chain into our analysis. To obtain paired TCRα/β sequences we performed scRNA-seq of SF CD4^+^ T cells from three patients (Supplemental Table 3). Clones that could be linked to the ‘specificity cluster’ by TCRβ specificity groups revealed a significant enrichment towards usage of the gene segment TRAV23/DV6 compared to all other clones which rather displayed random usage of TRAV segments (64% vs. 4.7% of the clones respectively, p< 0.0001 by Fisher’s exact test; Figure 3D and Supplemental Figure 5A). This significant enrichment of TRAV23/DV6 usage was also observed when filtering the clones for the above delineated surrogate marker combinations of the TCRβ chain (Figure 3E and Supplemental Figure 5B, C). Notably, whereas the CDR3β motive alone identified clones that in 52-62% also used TRAV23/DV6, combination of the CDR1β and CDR3β motive increased this frequency to 67-89% without substantial loss of identified clone numbers (Figure 3E). Hence, the combination of the CDR3β motive (’SL’ or ’SV’ at IMGT positions 111 and 112) with the CDR1β motive (’GH’ at IMGT positions 28 and 29), hereafter termed the ’ARLA motive,’ demonstrated the highest specificity without comprising sensitivity as a surrogate marker for identifying T cell clones that make up the identified ’specificity cluster’ in ARLA patients.

To investigate the specificity of the ‘ARLA motive’, we explored the CD4^+^ TCR repertoire within various disease conditions for its presence. Initially, we assessed published TCRβ sequences known to target microbial antigens or autoantigens for the defined surrogate markers (n=2094 from VDJdb (24), n=12 from (25)). The distinct ARLA-associated CDR3β motive was found in a few sequences, however, the specific combination of this motive together with the distinct VJ combination was not detected in any of the sequences (Supplemental Figure 6; CDR1β motive frequencies could not be assessed due to missing information in databases). The CDR3β motive was similarly low in the SF CD4^+^ TCR repertoire from two North American patients with LA and could not be identified in published TCRβ sequences from HLA-DRB1*04:01 restricted OspA_163-174_-specific T cell clones (Supplemental Figure 6B and 7) (25, 26). Additionally, the ‘ARLA motive’ was almost absent in the SF CD4^+^ T cell repertoire in patients with JIA and rheumatoid arthritis (RA) (Supplemental Figure 7) (26–28).

We next extended our analysis from the five patients with at least one HLA-DRB1*11 allele to all 12 ARLA patients with available TCR sequences and performed GLIPH2 on TCRβ sequences derived from bulk sorted PD-1^hi^CD4^+^ T cells from all ARLA patients as well as on published data from RA or JIA patients (input: 20,486, 25,095 and 8,698 sequences from ARLA, JIA and RA respectively; (22, 27)) and visualized the results using network analysis. Remarkably, a cluster was identified almost exclusively composed of sequences from ARLA cells (93.6% of TCR sequences from ARLA, 96.4% of those from ARLA patients with HLA-DRB1*11), with no such ’private’ clusters existing for the other two diseases (Figure 4A). The majority of clones in this cluster exhibited TCRβ sequences matching the ‘ARLA motive’ (Figure 4B). To consider the patients’ genetic background, we compared TCRβ repertoires from SF PD-1^hi^HLA-DR^+^ CD4^+^ T cells of ARLA and JIA patients with known HLA-DRB1 genotypes. The ‘ARLA motive’ was only present in ARLA patients carrying HLA-DRB1*11 and HLA-DRB1*13 alleles. These alleles could be distinguished from all other HLA-DRB1 alleles present in the cohort by the presence of serine at position 13 (Ser13), which is known to influence specific peptide binding properties and shape TCR selection (Supplemental Figure 8) (29, 30). The patient groups did not differ in the general distribution of the distinct TCRβ V or J genes used by the ‘ARLA motive’ (Supplemental Figure 9). However, the distinct ‘ARLA motive’ was significantly enriched in HLA-DRB1 Ser13 positive ARLA patients compared to ARLA patients without Ser13 alleles and JIA patients, irrespective of the HLA-DRB1 background (Figure 4C). This observation could be replicated in another dataset focusing on total CD4^+^ T cells derived from SF of JIA and ARLA patients (Supplemental Figure 10) (26).

**Figure 4.**
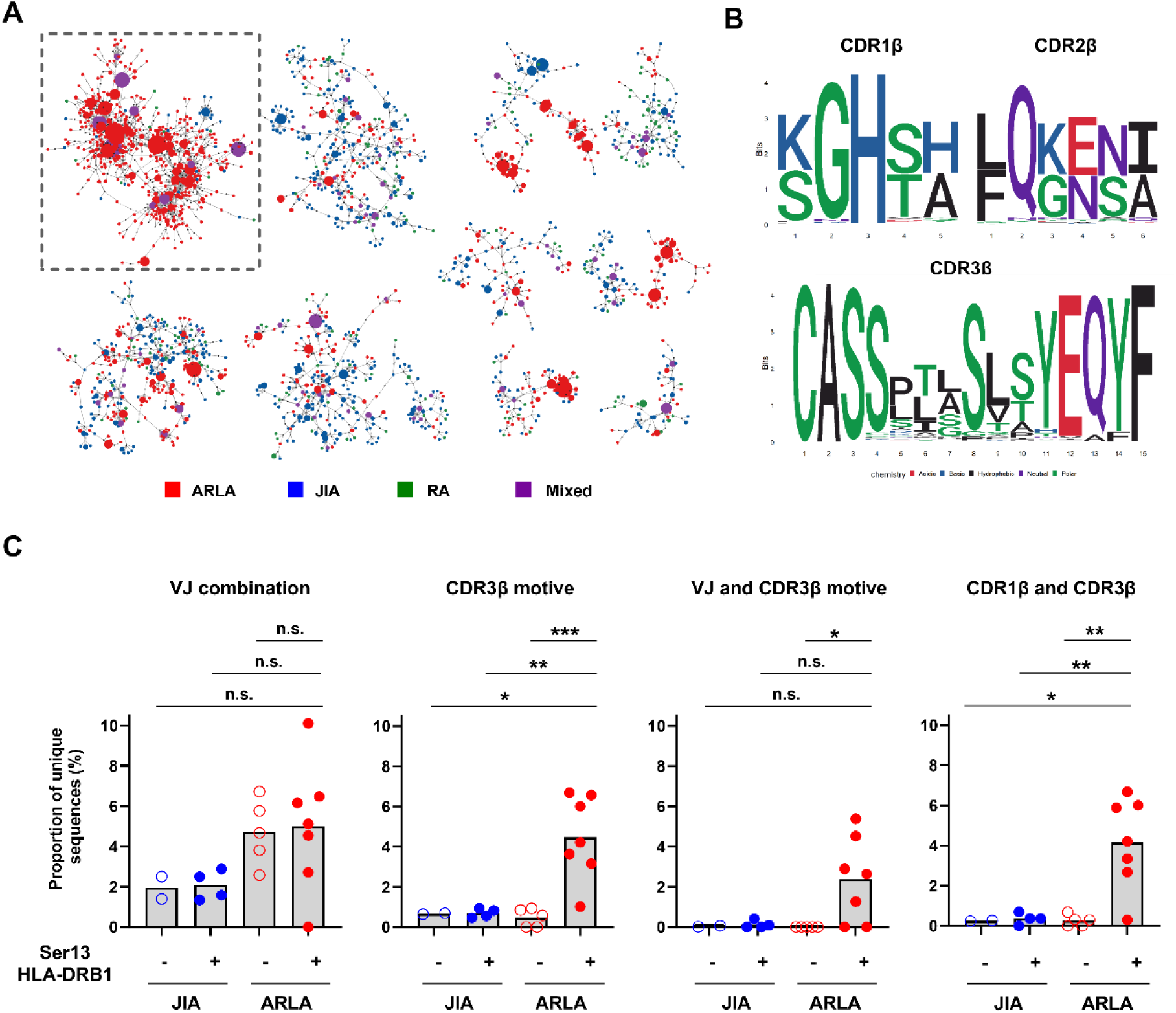
– Surrogate markers for ARLA-associated TCRs are disease specific and HLA-DRB1 restricted. **(A)** Network representation of TCR specificity groups enriched by GLIPH2 in synovial fluid (SF) PD-1^hi^CD4^+^ T cells from twelve ARLA, six JIA and three RA patients. Only specificity groups containing sequences from multiple patients are included and only networks with at least 50 members are shown. Motives are represented by small black circles and corresponding CDR3 sequences by colored circles. The circle sizes reflect the absolute abundances of unique CDR3 amino acid (aa) sequences across all patients. **(B)** Sequence plots showcasing the aa sequences in CDR1-3β, derived from sequences within the highlighted network on the left, are displayed. To generate these sequence plots, sequences were filtered for the most abundant length of each CDR. **(C)** Frequencies of indicated surrogate markers in TCRβ sequences of FACS sorted SF PD-1^hi^HLA-DR^+^CD4+ cells from children with JIA (n=6) and ARLA (n=12) determined by bulk sequencing. Patients exhibiting Serine at position 13 (Ser13) of HLA-DRB1 on at least one allele are denoted by filled circles. One-way ANOVA and multiple comparisons (to ‘ARLA +’) corrected with Dunnetts formula; n.s.: p>0.05, *: p<0.05, **: p<0.01, ***: p<0.001.

Hence, focusing on ARLA patients with a distinct HLA-DRB1 background allowed us to identify a TCRβ motive that served as a surrogate marker for the identification of ARLA-specific TCRs.

### Clonally expanded peripheral T helper cells dominate the CD4^+^ T cell landscape in the joints of patients with ARLA

To link TCR specificity to function, we next aimed to analyze the phenotype of CD4^+^ T cells expressing ARLA-specific TCRs in more detail. For this approach, we first comprehensively characterized the phenotype, functional state and clonality of SF Th cells in ARLA by conducting an in-depth investigation using combined scRNA-seq and single-cell TCR sequencing (scTCR-seq; 10X Genomics) of sorted CD4^+^ T cells obtained from the SF of three ARLA patients (Supplemental Table 3). Following demultiplexing, quality control and data integration, we successfully identified and analyzed a total of 12,622 CD4^+^ T cells for transcriptomic analysis.

By performing clustering analysis on the integrated transcriptomic data, we identified seven distinct clusters within the SF CD4^+^ T cells (Figure 5A). The cluster were equally represented between the three patients and TCR sequencing revealed sufficient coverage (Supplemental Figure 11). Cluster 0 emerged as the dominant cluster, exhibiting transcriptional similarities with cluster 3. Remarkably, these two clusters constituted approximately half of the total T cells in each patient (Figure 5B). Both clusters displayed heightened expression of *HLA-DR*, *PDCD1* (encoding PD-1) and *IL21* (encoding IL-21). Additionally, we observed upregulation of *LAG3*, *ALOX5AP* and *KLRB1* (encoding CD161) in both clusters, which together are well-established transcriptomic markers of peripheral T helper (T_PH_) cells (Figure 5C, Supplemental Figure 12 and Supplemental Table 4). Despite their similarities, only a few genes showed differential expression between the two T_PH_ cell clusters, with *CXCL13* being the most significant (Supplemental Table 5). This finding led us to classify cluster 0 and cluster 3 as CXCL13^low^ T_PH_ cells and CXCL13^high^ T_PH_ cells, respectively. The characteristic chemokine/cytokine pattern of the T_PH_ cluster with expression of CXCL13, IL-21 and IFN-γ could be recapitulated on protein level using flow cytometry (Figure 5D). Cluster 1 exhibited a significant enrichment in the expression of naïve T cell markers (*CCR7*, *SELL* and *TCF7*) as well as central memory marker genes (*IL7R*, *EEF1A1*) resulting in classification of cluster 1 as a “naïve/resting memory" T cell cluster. In contrast, cluster 2 showed heightened expression of cytotoxic marker genes but also immune regulatory genes (*IL10)*, while cluster 6 displayed an abundance of marker genes associated to regulatory T cells (Treg) (Figure 5C, Supplemental Figure 12 and Supplemental Table 4). As a result of these distinctive expression profiles, we labeled cluster 2 as a “cytotoxic/T regulatory 1 (Tr1) cluster”, and cluster 6 as a “Treg cluster”. Cluster 4 exhibited differential expression of mitochondrial genes and genes previously reported to be expressed in "Humanin" and "SESN3" CD4^+^ T cells in the SF of RA patients (31). Finally, the remaining cells in cluster 5 were predominantly characterized by marker genes associated with proliferating CD4^+^ T cells, indicating an actively dividing subpopulation within the synovial T cell pool (Figure 5C, Supplemental Figure 12 and Supplemental Table 4).

**Figure 5.**
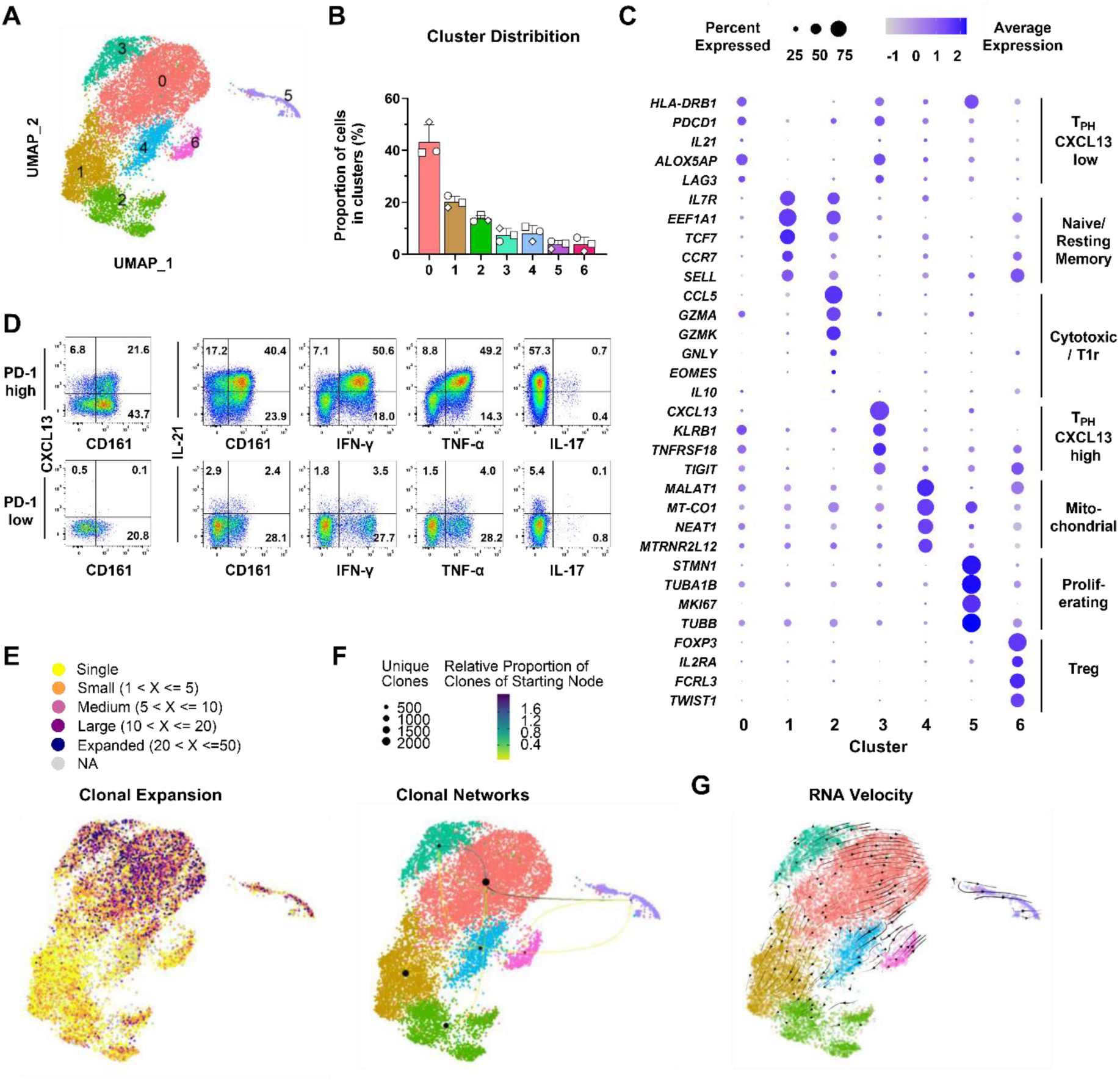
– The synovial CD4^+^ T cell landscape in ARLA is dominated by clonally expanded peripheral T helper cells. **(A)** UMAP representations of cell clustering via scRNA-seq analysis of synovial fluid (SF) CD4^+^ T cells from three ARLA patients. **(B)** Mean frequency of cells allocated to the respective clusters. Symbols represent individual patients, error bars indicate standard deviation. **(C)** 2-D dot plot showing the expression of selected marker genes. The area of the dots indicates the percentage of cells within the cluster expressing the gene, the color represents the average expression level. **(D)** Representative dot blots showing cytokine expression in SF PD-1^hi^ (upper row) and PD-1^lo/-^ CD4^+^ cells, assessed by flow cytometry. **(E)** Clonal expansion is depicted based on the size of individual clones, determined through paired TCRα/β sequencing, represented on the UMAP plot from panel A.**(F)** Clonal connectivities between individual clusters are illustrated by arrows, with darker colors indicating a higher proportion of clones originating from the starting cluster. **(G)** Possible developmental trajectories projected onto the UMAP representation from panel A, inferred by RNA velocity analysis.

To gain further insights into the tissue-specific clonal expansion and activation of each T cell cluster, we conducted an analysis of the TCR usage among all T cells that exhibited a productive TCRβ sequence. By integrating the scTCR-seq data into the analysis, we observed the most significant clonal expansion within the two T_PH_ clusters (Figure 5E and Supplemental Figure 13A). Notably, the largest clonal overlap was observed between these two T_PH_ clusters and the cluster of proliferating cells (Figure 5F and Supplemental Figure 13B), suggesting a potential shared developmental trajectory among these three clusters. In contrast, the overlap between the T_PH_ clusters and the two regulatory cluster (Tr1, cluster 2 and Treg, cluster 6) was minimal. RNA velocity analysis supported this observation and could not reveal a trajectory between the T_PH_ clusters and the other effector T cell clusters (Figure 5G).

In conclusion, these findings characterize SF T_PH_ cells as the dominant clonally expanding cell population suggesting that these cell subset might reflect the ongoing T helper response in the inflamed joints of patients with ARLA.

### T cells with ARLA specific TCRs reside in T_PH_ clusters and show signs of T cell receptor driven activation

Having identified surrogate markers for disease-specific TCRs in ARLA patients, we finally aimed at connecting TCR specificity with T cell phenotype and function. To achieve this, we mined the combined scRNA-seq/scTCR-seq data set of the three ARLA patients (each of them carrying at least one HLA-DRB1*11 allele) for T cell clones that displayed the TCRβ ‘ARLA motive’ (CDR1β and CDR3β). Out of 9,344 total cells, we identified 158 cells harboring this motive. To distinguish cells with TCR specificities unrelated to ARLA and potentially activated by bystander activation, we used TCRβ sequences with known specificities against viral antigens independent of a matching HLA background from the public VDJdb database to infer "viral motives" through the GLIPH2 algorithm (Supplemental Table 6) (24). We then searched the scRNA-seq/scTCR-seq dataset from ARLA patients for these "viral motives," resulting in a group of 625 cells.

Our analysis, applying these surrogate markers, revealed that the majority of cells with TCRs containing the ‘ARLA motive’ resided in the T_PH_ cell clusters 0 and 3 and marginally in proliferating cluster 5 (Figure 6A and B). However, they were almost absent in the Treg cluster 6 as well as in the Tr1 cluster 2. Additionally, when assessing the relative enrichment of the most frequent aa doublets (> 1% of doublets) at certain positions in the CDR3β in an unbiased manner, the only enrichment above background was observed for the doublet ’SL’ at CDR3β 111/112 (which constitutes to the ARLA CDR3β motive) in the T_PH_ clusters and proliferating cluster (Supplemental Figure 14). In contrast, T cells expressing TCRs with ‘viral motives’ were randomly distributed across the seven clusters, resembling the distribution of T cells not assigned to either TCR motive group (Figure 6A and B). Consistent with this observation, the proportion of cells in expanding clones was significantly higher in the ARLA group, and among them, the frequency of convergent clones appeared to be higher in cells with ‘ARLA motive’ (Supplemental Figure 15A and B).

**Figure 6.**
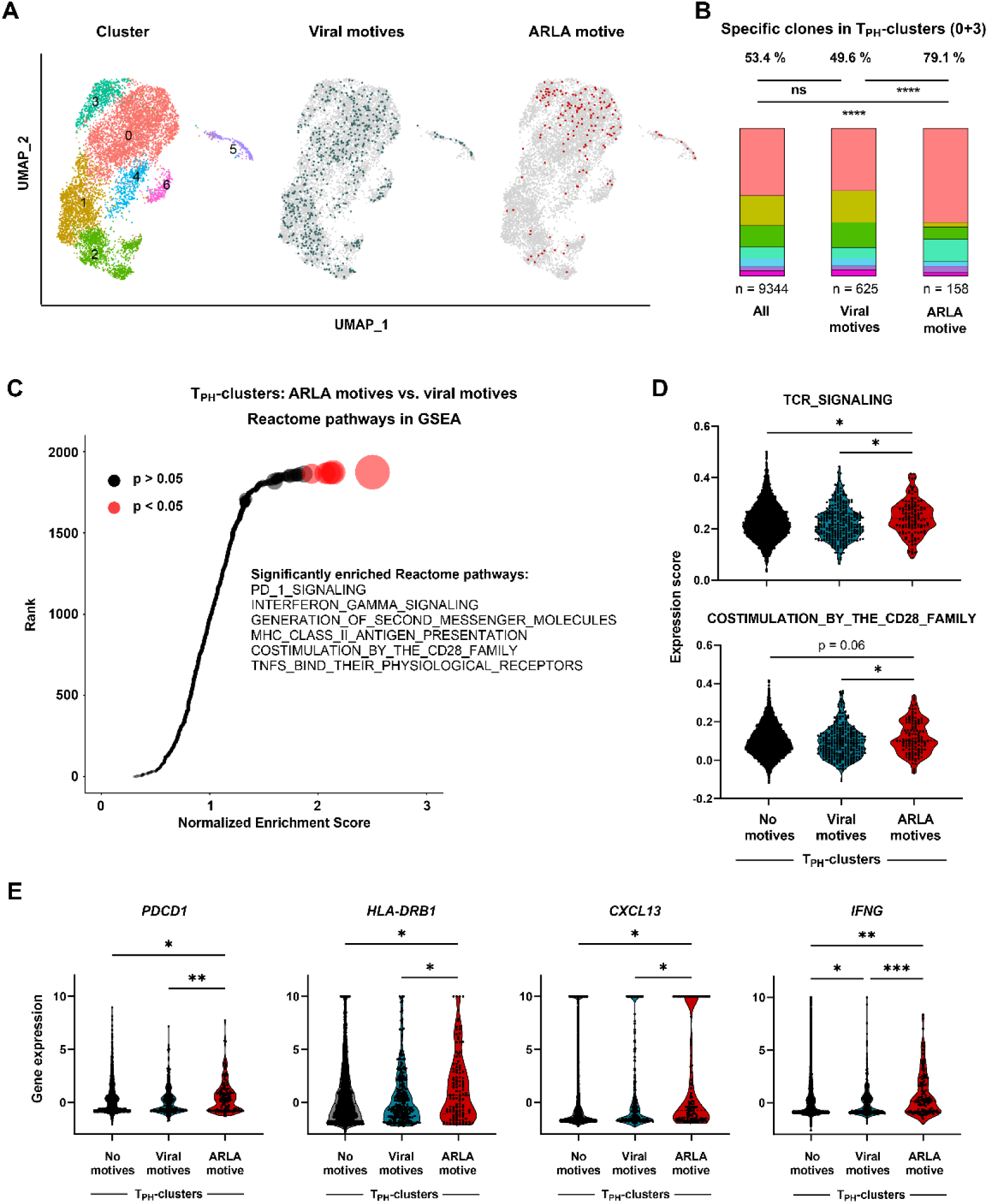
– ARLA-specific T cell clones map to the peripheral T helper cluster and show signs of T cell receptor driven activation. **(A)** UMAP representations of cell clustering via scRNA-seq analysis of synovial fluid (SF) CD4^+^ T cells from three ARLA patients (from Fig. 5A). In the center, cells with ‘viral’ TCR motives are highlighted, while on the right those with the ARLA motive (CDR1β and CDR3β motive) are highlighted. **(B)** Relative distribution of cells within each cluster, categorized by color as depicted in panel A, considering all cells or cells containing either viral or ARLA TCR motives. Fisher’s exact test between indicated groups; n.s.: p>0.05, ****: p<0.0001. **(C)** Normalized enrichment scores (NES) from Gene Set Enrichement Analysis (GSEA) analysis using Reactome pathways with differentially expressed genes between T_PH_ cells (cluster 0 and 3) with either ‘viral’ or ARLA TCR motives. Pathways are ranked based on their NES, with circle size corresponding to negative log10 (adjusted p value). Red circles represent pathways with an adjusted p value < 0.05. The significantly enriched pathways are identified by name. **(D)** Genes associated with TCR signaling-related GSEA pathways were used as input for the AddModuleScore function from Seurat. Resulting scores per cell are plotted and compared between T_PH_ cells containing no motives, ‘viral’ or ARLA TCR motives. **(E)** Expression of selected activation and effector genes in the same groups as in D. One-way ANOVA with Tukey’s multiple comparisons test; p values < 0.1 are shown, *: p<0.05, **: p<0.01,***: p<0.001.

Subsequently, we investigated the transcriptome of ARLA-specific clones to understand their modes of cellular activation. To reduce a bias by cells with ‘viral’ TCR motives from non-T_PH_ clusters, we exclusively focused the analysis on cells from the T_PH_ clusters (0 and 3). By applying differentially expressed genes between T_PH_ cells with ‘ARLA motives’ and ‘viral motives’ to GSEA Reactome pathway analysis, we found a significantly increased normalized enrichment score for pathways indicating signaling via the TCR and costimulatory receptors in the ‘ARLA motive’ group (Figure 6C). Additionally, creating scores applicable to single-cell gene expression data by utilizing genes from GSEA pathways for TCR signaling and costimulatory signaling resulted in significantly higher scores for T_PH_ cells in the ARLA motive group compared to both the group of T_PH_ cells with viral motives and that of T_PH_ cells lacking any of these motives (Figure 6D). Hence, even within the T_PH_ clusters those T cells expressing ARLA-specific TCR motives showed increased signs of continuous antigen engagement.

Finally, we assessed the effector programs of T_PH_ cells with ARLA-specific TCRs by comparing the expression of selected effector genes within these cells to T_PH_ cells expressing TCRs with no or ‘viral’ motives. Despite using exclusively T_PH_ cells as controls, genes associated with the T_PH_ phenotype (*PDCD1*, *HLA-DRB1*) and function (*CXCL13*) showed even higher expression in the group with ‘ARLA motives’ (Figure 6E). Additionally, T_PH_ cells bearing the TCR ‘ARLA motive’ also displayed higher expression of *IFNG* indicating an upregulated pro-inflammatory effector program in these cells (Figure 6E).

In conclusion, within the inflamed joints of ARLA patients, we could track T cells with disease-specific TCR that display signs of ongoing TCR triggering and express a pathogenic effector program resembling T_PH_ cells.

## DISCUSSION

By employing high-throughput TCR sequencing in conjunction with scRNA-seq, we have identified surrogate markers associated with disease-specific TCRs in postinfectious LA. These markers can be harnessed to dissect the pathogenic T cell response in the inflamed joints of patients. Our findings provide evidence that disease-specific T cells are the driving force behind the expansion of a dominant T_PH_ cell population within the joints of ARLA patients. These cells exhibit sustained proliferation and replenishment throughout the course of the disease, likely in response to antigen-driven TCR signaling. Notably, ARLA-specific T_PH_ cells display a pathogenic effector program with heightened expression of IFN-γ as well as CXCL13, which have dominant roles in tissue inflammation and tertiary lymphoid structure formation.

Thus far, presumed targets of the pathological T-cell response in ARLA encompass a wide spectrum of antigens, ranging from *Borrelia* components (e.g. OspA) to autoantigens associated to vascular tissue (ECGF, annexin A2 and apoB-100) or extracellular matrix (fibronectin-1, laminin B2 and collagen) (14–17, 25, 32, 33). These diverse T cell responses have been integrated into a framework indicating that exaggerated immune reactions to OspA might disrupt T cell tolerance, triggering epitope spreading from *Borrelia* antigens to autoantigens like extracellular matrix proteins (7). Notably, T cell responses against these autoantigens have been specifically detected in ARLA patients using T-cell activation assays or peptide-MHC tetramers. The use of these techniques exhibits exceptional sensitivity in discerning antigen-specific T cells contingent upon the knowledge of disease-relevant HLA alleles and their associated epitopes. As a further limitation, activation assays can induce significant alterations in antigen-reactive cells, leading to a bias in their phenotypic and functional characterization. Given the ambiguity encompassing the dominant direction of T cell responses in postinfectious ARLA outside North American (19, 20), accurately and comprehensively predicting the primary targets of the pathogenic T cell response might also prove challenging when employing candidate antigen approaches. In order to overcome these challenges, we decided to use an unbiased approach and employed the GLIPH2 algorithm for the clustering of TCRs that exhibit recognition of the same epitope predicated by shared structural similarity within the CDR3 (23). This algorithm has gained widespread utilization for the detection of antigen-specific TCRs in instances where information pertaining to the antigens is not readily available (34). Additionally, by combining TCR sequence information with gene expression data we could link TCR specificity to cellular function. By this approach we are able to provide a dataset that may facilitate the comprehensive and unbiased identification of target antigens of disease-associated TCRs.

In the synovial CD4^+^ T cell population present in the joints of ARLA patients, we observed a significant increase in activated effector T cells, characterized by heightened expression of PD-1 and HLA-DR, suggesting recent antigen-induced activation. By leveraging this particular phenotype to enrich antigen-reactive T cells and employing high-throughput sequencing, we were able to identify shared disease-specific TCR motives among patients, which were HLA-DRB1-restricted. These motives were primarily determined by a non-germline encoded SV/SL at a conserved position 111/112 within the CDR3β and the utilization of V segments containing aa GH at IMGT position 28/29 (CDR1β), along with the usage of TRAV23/DV6 in the TCRα, without the identification of a specific motive therein. The canonical CDR1/3β sequence functioned as a reliable surrogate marker for identification of respective TCRs. A similar structural configuration, characterized by the preferential Vβ/Jβ usage, a non-germline encoded CDR3β motive and biased *TRAV* pairing, was identified in gluten-specific, HLA-restricted CD4^+^ T cell clones in celiac disease (35). Here, the non-germline CDR3β motive made crucial interactions with the gliadin epitope as well as the HLA-DQ2.5 allele, thereby acting as a lynchpin. In ARLA, the marked diversity in nucleotide-level sequences encoding CDR3β motives, its persistent correlation with T cells throughout the course of the disease, and the partial replacement of TCR motive-bearing T cells by distinct clonal populations over time collectively align with the concept of a convergent T cell response instigated by local antigens. This suggestion finds further support in the observed HLA restriction, predominantly linked to HLA-DRB1*11 and HLA-DRB1*13, presumably dictated by the presence of specific aa at a discrete positions within the peptide-binding pocket 4 of the HLA molecule (HLA-DRB1 Ser13) (29). HLA-DRB1 position 13 has a strong association with CDR3β aa composition such being involved in shaping the TCR repertoire and mediates risk for multiple autoimmune diseases (30, 36).

Unexpectedly, our investigation revealed a notably high prevalence of the HLA-DRB1*11 allele within the cohort of ARLA patients under scrutiny. This particular allele had previously been identified as a potential protective factor against the development of an antibiotic-refractory disease course in a North American patient study (8). The cohort analyzed in our research comprises ARLA patients referred to our center from diverse regions, providing a representative snapshot - albeit with a limited number of participants - of the spectrum of affected children and adolescents in the German area. The distribution of *Borrelia* species and strains in this geographical region shows a notable difference compared to North America. In Germany, *B. afzelii* constituted approximately 60% of the infected ticks, followed by *B. garinii* and *B. burgdorferi s.s*., each accounting for around 10% (37). In contrast, *B. burgdorferi s.s.* stands as the prevailing species in North America (1). Despite this marked difference, it is generally believed that *B. burgdorferi s.s.* also remains the predominant species in cases of clinically evident LA in Europe (7). This, along with the prevalence of less inflammatory strains of *B. burgdorferi s.s*., might account for the lower occurrence and reduced disease severity of LA in Europe. Nonetheless, *B. afzelii* and *B. garinii* could each be detected in up to one third of European patients with LA (38, 39). Hence, it is reasonable to suggest that the European situation, involving LA, differs not only in the presence of diverse *B. burgdorferi s.s.* strains but also in the distribution of *Borrelia* species when juxtaposed with the North American context. Moreover, this disparity might extend to ARLA since the distribution of *Borrelia* species in a group of European patients with prolonged course of arthritis despite antibiotic treatment was also diverse and not restricted to *B. burgdorferi s.s*. (38). It is therefore reasonable to assume that, in comparison to the North American situation, different immunodominant *Borrelia* peptides and, consequently, different HLA-DRB1 alleles are involved in the pathogenesis of ARLA in Europe. Our elucidation of TCR motives within the context of HLA-DRB1*11-associated ARLA will facilitate the prospect of discerning putative target antigens linked to the pathogenic T-cell response in these afflicted individuals. This will also enable a comparative analysis with the antigens predominantly associated with known HLA-DRB1 risk alleles within the North American context.

Unsurprisingly, expansion of effector T cells bearing the presumed ARLA-specific TCR motive could not be documented in RA, in which the HLA-DRB1 Ser13 alleles are protective (36). In contrast, HLA-DRB Ser13 alleles were confirmed to predispose to JIA which phenotypically may resemble ARLA (40). Of note, the TCRβ repertoire of activated effector cells in the joints of HLA-DRB Ser13 positive JIA patients without serological evidence of prior *Borrelia* infection was distinct from that of ARLA patients with absence of the TCR ‘ARLA motive’. Hence, TCRs bearing the ‘ARLA motive’ may either recognize *Borrelia* antigens not present in the joint of JIA patients or more likely recognize autoantigens that are not targeted by the T cell response in JIA. Cumulatively, these observations substantiate the hypothesis that the dominate T cell response within the joints of ARLA patients is private and differs in its dominant antigen targets from other rheumatic diseases.

Our study was limited in its ability to determine whether the TCRs specifically identified in ARLA patients are also involved in the initiation of the pathogenic T cell response. While we successfully characterized the T cell response throughout the chronic phase of arthritis, we did not have access to samples from the initial disease onset of patients who later developed ARLA. Although these limitations constrain interpretation, the convergent T cell response observed in ARLA patients during the late disease course would align well with the proposed epitope spreading model (7, 14). The observation that T cells carrying TCRs with known specificities against viral antigens were randomly distributed among various T cell clusters and not enriched in the expanded T_PH_ effector cell cluster also refutes bystander activation as the primary mechanism by which T cells are recruited within the local T cell response.

The synovial transcriptomic landscape in patients with ARLA is characterized by a pronounced IFN-γ expression signature and high frequencies of IFN-y secreting T cells are present in SF of these patients (41–43). Furthermore, autoreactive T cells, identified through peptide-MHC tetramers recognizing extracellular matrix proteins, are notably enriched in T-bet-expressing cells (14). This observation implies that the pathogenic T cell response in ARLA is mediated by Th-1 cells. Our study refines this assumption by demonstrating that ARLA-specific T cells exhibit elevated levels of IFN-γ expression but can be more accurately classified as T_PH_ cells rather than classical Th-1 cells. T_PH_ cells are pivotal drivers of disease pathogenesis in numerous autoimmune conditions and are expanded in inflamed tissues of patients with RA, JIA and systemic lupus erythematosus (SLE) (22, 44, 45). The phenotype and function of T_PH_ cells exhibit partial overlap with follicular T helper cells, expressing B cell-supporting cytokines such as IL-21 and CXCL13 (46). However, they rather possess a chemokine receptor profile facilitating their accumulation at inflamed sites, along with Th-1-like features including T-bet, IFN-γ and TNF-α expression. Remarkably, T cells bearing the ‘ARLA motive’ were almost undetectable within the local Treg population as well as in the Tr1 cluster. These findings reinforce earlier observations of a disturbed Treg compartment in ARLA, indicating a lack of recruitment of T cells with regulatory functions into the local T cell response (47, 48). Our findings establish T_PH_ cells as the dominant population among synovial T helper cells in ARLA patients and link the convergent TCR response of ARLA-specific T cells to the functional profile of these cells.

In summary, we detected disease-specific TCRs within the inflamed joints of European children and adolescents with ARLA which will facilitate the identification of major antigen targets in this diseases. The ARLA-specific T cells displayed continuous proliferation, potentially triggered by localized TCR signaling, and demonstrated a pathogenic effector profile akin to peripheral T_PH_ cells, characterized by the expression of CXCL13 and IFN-γ. Consequently, T_PH_ cells should be acknowledged as the primary pathogenic Th cell subset in the inflamed joints of individuals with ARLA.

*TRBV* and *TRBJ* gene usage, along with the CDR3 amino acid (aa) sequences of the three most prevalent clones within each specificity group comprising the ’specificity cluster’ illustrated in Figure 2B. The aa motives characterizing each specificity group are summarized in the ’Pattern’ column. Sequences were filtered to include enriched patterns consisting of more than 20 unique CDR3s. In the CDR3 aa sequence, small capitals denote non-germline encoded aa. The total count of unique CDR3s forming the specificity groups, as well as the distribution of the sequences in the respective group and from the three most frequent ‘clones’ across the five analyzed patients are shown.

## METHODS

### Patients

Sex as a biological variable: Our study examined male and female patients, and similar findings are reported for both sexes.

Pediatric patients diagnosed with ARLA and JIA (as disease control) undergoing joint puncture for intra-articular steroid injection or diagnostic purposes were enrolled in this study (Supplemental Tables 1 and 3). ARLA was defined as arthritis with serological evidence of late-stage *Borrelia burgdorferi* infection (*Borrelia burgdorferi* IgG ELISA positive, IgG immunoblot with ≥ 5 positive bands; recomWell Borrelia IgG and IgM, and recomLine Borrelia IgG and IgM from Mikrogen Diagnostik, Neuried, Germany) that did not respond to at least two cycles of antibiotic treatment, including a combination of four weeks of amoxicillin, and/or four weeks of doxycycline, and/or two weeks of cefotaxime/ceftriaxone. SF samples from ARLA patients were obtained after completion of antibiotic treatment. JIA was defined according to the ILAR classification criteria and only JIA patients without serological evidence of prior *Borrelia* infection (IgG- and IgM-negative by ELISA) were included in this study as disease controls. The data for the JIA disease control group was partially retrieved from a previous study (22). HLA-typing was performed on DNA extracted from peripheral blood or buccal swabs by an Illumina MiSeq based next generation sequencing method at the DKMS Life Science Lab (Dresden, Germany). Patients were monitored at the University Children’s Hospital Würzburg, University Children’s Hospital Leipzig, and Children’s Hospital, Vivantes Klinikum im Friedrichshain, Berlin.

### Sample preparation

Mononuclear cells were isolated from SF using Ficoll density-gradient centrifugation. Mononuclear cells from synovial tissue were acquired through mechanical disaggregation, followed by Ficoll density-gradient centrifugation. Cells were preserved in 10% Fetal Calf Serum (FCS) 10% (Dimethylsulfoxid) DMSO 10% and stored in liquid nitrogen until further use.

### Immunohistochemistry

Synovia was fixed in 4% paraformaldehyde and embedded in paraffin after dehydration in alcohol. Immunohistochemistry was performed using standard diagnostic protocols.

### Antibodies, flow cytometry and cell sorting

Antibodies used for flow cytometry and cell sorting are listed in the supplemental methods. Mononuclear cells were stained in 1XPBS 0.5% bovine serum albumin (BSA) with appropriate antibodies at 4°C for 30 minutes. Intracellular staining was carried out according to the manufacturer’s instruction using Intracellular Fixation & Permeabilization Buffer (eBioscience). For detection of intracellular cytokine expression, mononuclear cells were stimulated with phorbol 12-myristate 13-acetate (PMA, 50 ng/ml; Sigma-Aldrich) and ionomycin (1 μg/ml; Sigma-Aldrich) with addition of Brefeldin A (5 μg/ml, BioLegend) for 4 hours before staining. For detection of intracellular CXCL13 expression, mononuclear cells were stimulated with CD3/CD28 Dynabeads (Thermo Fisher Scientific) at a bead to T cell ratio of 1:1 for 6 hours with addition of Brefeldin A for the last 4 hours. Flow cytometry data was acquired on a FACSCanto II (BD Biosciences) and analyzed with FlowJo version 10 (Tree Star). Cell sorting was performed on a FACSAria III (BD Biosciences). Alternatively, CD4^+^ T cells were purified using the EasySep™ Human CD4^+^ T Cell Isolation Kit (Stemcell Technologies, Vancouver, Canada).

### TCRβ repertoire sequencing

RNA from CD4^+^CD45RO^+^PD-1^hi^HLA-DR^+^ and CD4^+^CD45RO^+^PD-1^lo/-^HLA-DR^-^ sorted T cells obtained from SF and synovial tissue was purified using RNeasy Micro Kit (Qiagen). Total numbers of sorted cells from the two T cell populations were adjusted to the lowest cell number obtained within each patient. TCRβ repertoire sequencing was performed either by amplicon rescue multiplex PCR from a commercial provider (iRepertoire®, Huntsville, AL, USA) or by multiplex PCR using an in-house pipeline and sequenced by next generation sequencing on a MiSeq platform. Experimental details are described in the Supplemental Methods.

### TCRβ repertoire analysis

Resulting sequence data was analyzed using IMGT/HighV-QUEST, ARGalaxy, Alakazam, Immunarch and GLIPH2 (Grouping of Lymphocyte Interactions by Paratope Hotspots, version 2). Details of the analytical work-flows are described in the Supplemental Methods.

### 10x Genomics Chromium single-cell RNA-seq

SF CD4^+^ T cells from three ARLA patients (ARLA01, 02 and 05) were subjected to scRNA-seq (10x Genomics). For ARLA01 and 05 samples were multiplexed with TotalSeqC Hashtag Oligos (Biolegend; ARLA01: GTCAACTCTTTAGCG, ARLA05: TTCCGCCTCTCTTTG) prior to preparation of single cell suspensions. After encapsulation and barcoding (10x Genomics), cells were lysed and cDNA prepared to create a 5′ gene expression library and a VDJ gene-enriched library for TCR repertoire analysis. Libraries were sequenced using Illumina NovaSeq 6000. Raw data were processed by CellRanger (10x Genomics) with standard settings. Resulting output files were imported in R version 4.2.1 using the Read10X function of Seurat version 4.1.1 (49). Details of the further analytical work-flows are described in the Supplemental Methods.

### Adaption of published TCRβ data

Publicly available TCRβ repertoire sequencing data sets were re-analyzed for this study. The analytical work-flow is described in detail in the Supplemental Methods.

### Data visualization

Various R packages (ggplot2, ggalluvial, alakazam, immunarch, ggseqlogo, Seurat, scRepertoire, circlize) were employed for figure generation regarding TCRβ and scRNA-seq analysis. Additionally, Prism 10.1.0 (GraphPad) was utilized to visualize quantitative data generated from other sources.

### Statistics

Statistical analysis was performed using Prism 10.1.0 (GraphPad). Data are expressed as scattered individual values and mean ± standard deviation. Student’s t-test or One-way ANOVA with Dunnett’s multiple comparisons test were used for comparison of data sets with two or more continuous variables, respectively. For contingency tables chi-square with Yates’ correction was used. A p value <0.05 was considered as statistically significant.

### Study approval

Written informed consent was obtained from the legal guardians. The study was approved by the Research Ethics Committee of the University of Würzburg (299/17) and conducted in strict accordance with the principles of the Declaration of Helsinki.

## Data availability

Bulk TCR sequencing data is deposited in the NCBI Sequence Read Archive under BioProject ID PRJNA1054606. The single cell sequencing data discussed in this publication have been deposited in NCBI’s Gene Expression Omnibus and are accessible through GEO Series accession number GSE247675.

(https://www.ncbi.nlm.nih.gov/geo/query/acc.cgi?acc=GSE247675).

## ACKNOWLEDGEMENTS

We thank Gabriele Haase and Ursula Fischer for technical assistance. We thank the Core Unit for FACS of the IZKF Würzburg and the Single-Cell Center Würzburg for assistance with cell sorting and scRNA-seq. This works was supported by the German Research Foundation (MO 2160/4-1, HM). HM received funding from the Federal Ministry of Education and Research (BMBF; Advanced Clinician Scientist-Program INTERACT; 01EO2108) embedded in the Interdisciplinary Center for Clinical Research (IZKF) of the University Hospital Würzburg. JD was supported by the German Center for Infection Research (DZIF; Clinical Leave Program; TI07.001_007). JF was supported by the Interdisciplinary Center for Clinical Research (IZKF) Würzburg (Clinician Scientist Program, Z-2/CSP-30).

## AUTHOR CONTRIBUTIONS

JD and HM designed the study. JD, JF, JK and VB performed experiments. JD, JF, JK, AHW, CH, CK and HG collected data. JD, JF, JK, AHW, CH, CK, VB, HG, IC and FE analyzed data. JD and HM drafted the manuscript, and all authors revised and finally approved the manuscript.

## Notes

“The authors have declared that no conflict of interest exists.”

### Competing Interest Statement

The authors have declared no competing interest.

### Author Declarations

The Ethics Committee of the University of Wuerzburg gave ethical approval for this work.

## REFERENCES

1. Steere AC, Strle F, Wormser GP, Hu LT, Branda JA, Hovius JW, et al. Lyme borreliosis. Nat Rev Dis Primers. 2016;2:16090.

2. Singh SK, and Girschick HJ. Molecular survival strategies of the Lyme disease spirochete Borrelia burgdorferi. Lancet Infect Dis. 2004;4(9):575–83.

3. Piesman J, and Gern L. Lyme borreliosis in Europe and North America. Parasitology. 2004;129 Suppl:S191-220.

4. Arvikar SL, and Steere AC. Lyme Arthritis. Infect Dis Clin North Am. 2022;36(3):563–77.

5. Bentas W, Karch H, and Huppertz HI. Lyme arthritis in children and adolescents: outcome 12 months after initiation of antibiotic therapy. J Rheumatol. 2000;27(8):2025–30.

6. Horton DB, Taxter AJ, Davidow AL, Groh B, Sherry DD, and Rose CD. Pediatric Antibiotic-refractory Lyme Arthritis: A Multicenter Case-control Study. J Rheumatol. 2019;46(8):943–51.

7. Lochhead RB, Strle K, Arvikar SL, Weis JJ, and Steere AC. Lyme arthritis: linking infection, inflammation and autoimmunity. Nat Rev Rheumatol. 2021;17(8):449–61.

8. Steere AC, Klitz W, Drouin EE, Falk BA, Kwok WW, Nepom GT, et al. Antibiotic-refractory Lyme arthritis is associated with HLA-DR molecules that bind a Borrelia burgdorferi peptide. J Exp Med. 2006;203(4):961–71.

9. Kamradt T, Lengl-Janssen B, Strauss AF, Bansal G, and Steere AC. Dominant recognition of a Borrelia burgdorferi outer surface protein A peptide by T helper cells in patients with treatment-resistant Lyme arthritis. Infect Immun. 1996;64(4):1284–9.

10. Lengl-Janssen B, Strauss AF, Steere AC, and Kamradt T. The T helper cell response in Lyme arthritis: differential recognition of Borrelia burgdorferi outer surface protein A in patients with treatment-resistant or treatment-responsive Lyme arthritis. J Exp Med. 1994;180(6):2069–78.

11. Kalish RA, Leong JM, and Steere AC. Association of treatment-resistant chronic Lyme arthritis with HLA-DR4 and antibody reactivity to OspA and OspB of Borrelia burgdorferi. Infect Immun. 1993;61(7):2774–9.

12. Chen J, Field JA, Glickstein L, Molloy PJ, Huber BT, and Steere AC. Association of antibiotic treatment-resistant Lyme arthritis with T cell responses to dominant epitopes of outer surface protein A of Borrelia burgdorferi. Arthritis Rheum. 1999;42(9):1813–22.

13. Steere AC, Falk B, Drouin EE, Baxter-Lowe LA, Hammer J, and Nepom GT. Binding of outer surface protein A and human lymphocyte function-associated antigen 1 peptides to HLA-DR molecules associated with antibiotic treatment-resistant Lyme arthritis. Arthritis Rheum. 2003;48(2):534–40.

14. Kanjana K, Strle K, Lochhead RB, Pianta A, Mateyka LM, Wang Q, et al. Autoimmunity to synovial extracellular matrix proteins in patients with postinfectious Lyme arthritis. J Clin Invest. 2023;133(17).

15. Crowley JT, Strle K, Drouin EE, Pianta A, Arvikar SL, Wang Q, et al. Matrix metalloproteinase-10 is a target of T and B cell responses that correlate with synovial pathology in patients with antibiotic-refractory Lyme arthritis. J Autoimmun. 2016;69:24–37.

16. Pianta A, Drouin EE, Crowley JT, Arvikar S, Strle K, Costello CE, et al. Annexin A2 is a target of autoimmune T and B cell responses associated with synovial fibroblast proliferation in patients with antibiotic-refractory Lyme arthritis. Clin Immunol. 2015;160(2):336–41.

17. Crowley JT, Drouin EE, Pianta A, Strle K, Wang Q, Costello CE, et al. A Highly Expressed Human Protein, Apolipoprotein B-100, Serves as an Autoantigen in a Subgroup of Patients With Lyme Disease. J Infect Dis. 2015;212(11):1841–50.

18. Londono D, Cadavid D, Drouin EE, Strle K, McHugh G, Aversa JM, et al. Antibodies to endothelial cell growth factor and obliterative microvascular lesions in the synovium of patients with antibiotic-refractory lyme arthritis. Arthritis Rheumatol. 2014;66(8):2124–33.

19. Dressler F, Ackermann R, and Steere AC. Antibody responses to the three genomic groups of Borrelia burgdorferi in European Lyme borreliosis. J Infect Dis. 1994;169(2):313–8.

20. Li X, Strle K, Wang P, Acosta DI, McHugh GA, Sikand N, et al. Tick-specific borrelial antigens appear to be upregulated in American but not European patients with Lyme arthritis, a late manifestation of Lyme borreliosis. J Infect Dis. 2013;208(6):934–41.

21. Christophersen A, Lund EG, Snir O, Sola E, Kanduri C, Dahal-Koirala S, et al. Distinct phenotype of CD4(+) T cells driving celiac disease identified in multiple autoimmune conditions. Nat Med. 2019;25(5):734–7.

22. Fischer J, Dirks J, Klaussner J, Haase G, Holl-Wieden A, Hofmann C, et al. Effect of Clonally Expanded PD-1(high) CXCR5-CD4+ Peripheral T Helper Cells on B Cell Differentiation in the Joints of Patients With Antinuclear Antibody-Positive Juvenile Idiopathic Arthritis. Arthritis Rheumatol. 2022;74(1):150–62.

23. Huang H, Wang C, Rubelt F, Scriba TJ, and Davis MM. Analyzing the Mycobacterium tuberculosis immune response by T-cell receptor clustering with GLIPH2 and genome-wide antigen screening. Nat Biotechnol. 2020;38(10):1194–202.

24. Goncharov M, Bagaev D, Shcherbinin D, Zvyagin I, Bolotin D, Thomas PG, et al. VDJdb in the pandemic era: a compendium of T cell receptors specific for SARS-CoV-2. Nat Methods. 2022;19(9):1017–9.

25. Ausubel LJ, O’Connor KC, Baecher-Allen C, Trollmo C, Kessler B, Hekking B, et al. Characterization of in vivo expanded OspA-specific human T-cell clones. Clin Immunol. 2005;115(3):313–22.

26. Henderson LA, Volpi S, Frugoni F, Janssen E, Kim S, Sundel RP, et al. Next-Generation Sequencing Reveals Restriction and Clonotypic Expansion of Treg Cells in Juvenile Idiopathic Arthritis. Arthritis Rheumatol. 2016;68(7):1758–68.

27. Sakuragi T, Yamada H, Haraguchi A, Kai K, Fukushi JI, Ikemura S, et al. Autoreactivity of Peripheral Helper T Cells in the Joints of Rheumatoid Arthritis. J Immunol. 2021;206(9):2045–51.

28. Maschmeyer P, Heinz GA, Skopnik CM, Lutter L, Mazzoni A, Heinrich F, et al. Antigen-driven PD-1(+) TOX(+) BHLHE40(+) and PD-1(+) TOX(+) EOMES(+) T lymphocytes regulate juvenile idiopathic arthritis in situ. Eur J Immunol. 2021;51(4):915–29.

29. Bondinas GP, Moustakas AK, and Papadopoulos GK. The spectrum of HLA-DQ and HLA-DR alleles, 2006: a listing correlating sequence and structure with function. Immunogenetics. 2007;59(7):539–53.

30. Ishigaki K, Lagattuta KA, Luo Y, James EA, Buckner JH, and Raychaudhuri S. HLA autoimmune risk alleles restrict the hypervariable region of T cell receptors. Nat Genet. 2022;54(4):393–402.

31. Argyriou A, Wadsworth MH, 2nd, Lendvai A, Christensen SM, Hensvold AH, Gerstner C, et al. Single cell sequencing identifies clonally expanded synovial CD4(+) TPH cells expressing GPR56 in rheumatoid arthritis. Nat Commun. 2022;13(1):4046.

32. Drouin EE, Seward RJ, Strle K, McHugh G, Katchar K, Londono D, et al. A novel human autoantigen, endothelial cell growth factor, is a target of T and B cell responses in patients with Lyme disease. Arthritis Rheum. 2013;65(1):186–96.

33. Drouin EE, Glickstein L, Kwok WW, Nepom GT, and Steere AC. Searching for borrelial T cell epitopes associated with antibiotic-refractory Lyme arthritis. Mol Immunol. 2008;45(8):2323–32.

34. Greaves SA, Ravindran A, Santos RG, Chen L, Falta MT, Wang Y, et al. CD4+ T cells in the lungs of acute sarcoidosis patients recognize an Aspergillus nidulans epitope. J Exp Med. 2021;218(10).

35. Ciacchi L, Reid HH, and Rossjohn J. Structural bases of T cell antigen receptor recognition in celiac disease. Curr Opin Struct Biol. 2022;74:102349.

36. Raychaudhuri S, Sandor C, Stahl EA, Freudenberg J, Lee HS, Jia X, et al. Five amino acids in three HLA proteins explain most of the association between MHC and seropositive rheumatoid arthritis. Nat Genet. 2012;44(3):291–6.

37. Glass A, Springer A, Raulf MK, Fingerle V, and Strube C. 15-year Borrelia prevalence and species distribution monitoring in Ixodes ricinus/inopinatus populations in the city of Hanover, Germany. Ticks Tick Borne Dis. 2023;14(1):102074.

38. Grillon A, Scherlinger M, Boyer PH, De Martino S, Perdriger A, Blasquez A, et al. Characteristics and clinical outcomes after treatment of a national cohort of PCR-positive Lyme arthritis. Semin Arthritis Rheum. 2019;48(6):1105–12.

39. Corre C, Coiffier G, Le Goff B, Ferreyra M, Guennic X, Patrat-Delon S, et al. Lyme arthritis in Western Europe: a multicentre retrospective study. Eur J Clin Microbiol Infect Dis. 2022;41(1):21–7.

40. Hinks A, Bowes J, Cobb J, Ainsworth HC, Marion MC, Comeau ME, et al. Fine-mapping the MHC locus in juvenile idiopathic arthritis (JIA) reveals genetic heterogeneity corresponding to distinct adult inflammatory arthritic diseases. Ann Rheum Dis. 2017;76(4):765–72.

41. Gross DM, Steere AC, and Huber BT. T helper 1 response is dominant and localized to the synovial fluid in patients with Lyme arthritis. J Immunol. 1998;160(2):1022–8.

42. Lochhead RB, Arvikar SL, Aversa JM, Sadreyev RI, Strle K, and Steere AC. Robust interferon signature and suppressed tissue repair gene expression in synovial tissue from patients with postinfectious, Borrelia burgdorferi-induced Lyme arthritis. Cell Microbiol. 2019;21(2):e12954.

43. Lochhead RB, Ordonez D, Arvikar SL, Aversa JM, Oh LS, Heyworth B, et al. Interferon-gamma production in Lyme arthritis synovial tissue promotes differentiation of fibroblast-like synoviocytes into immune effector cells. Cell Microbiol. 2019;21(2):e12992.

44. Rao DA, Gurish MF, Marshall JL, Slowikowski K, Fonseka CY, Liu Y, et al. Pathologically expanded peripheral T helper cell subset drives B cells in rheumatoid arthritis. Nature. 2017;542(7639):110-4.

45. Bocharnikov AV, Keegan J, Wacleche VS, Cao Y, Fonseka CY, Wang G, et al. PD-1hiCXCR5-T peripheral helper cells promote B cell responses in lupus via MAF and IL-21. JCI Insight. 2019;4(20).

46. Yoshitomi H, and Ueno H. Shared and distinct roles of T peripheral helper and T follicular helper cells in human diseases. Cell Mol Immunol. 2021;18(3):523–7.

47. Shen S, Shin JJ, Strle K, McHugh G, Li X, Glickstein LJ, et al. Treg cell numbers and function in patients with antibiotic-refractory or antibiotic-responsive Lyme arthritis. Arthritis Rheum. 2010;62(7):2127–37.

48. Vudattu NK, Strle K, Steere AC, and Drouin EE. Dysregulation of CD4+CD25(high) T cells in the synovial fluid of patients with antibiotic-refractory Lyme arthritis. Arthritis Rheum. 2013;65(6):1643–53.

49. Hao Y, Hao S, Andersen-Nissen E, Mauck WM, 3rd, Zheng S, Butler A, et al. Integrated analysis of multimodal single-cell data. Cell. 2021;184(13):3573–87 e29.

