## Supplemental material for "Identification of disease-specific TCRs maintaining pathogenic T helper cell responses in postinfectious Lyme Arthritis"

**SUPPLEMENTAL DATA**

SUPPLEMENTAL METHODS

**Antibodies, flow cytometry and cell sorting**

The following antibodies were used for flow cytometry and cell sorting: CD3 BV510 (clone SK7), CD4 APC-Cy7 (clone OKT4), PD-1 PE-Cy7 (clone EH12.2H7), CD45RO PerCP-Cy5.5 (clone UCHL1), CD161 PerCP-Cy5.5 (clone HP-3G10), CXCR5 BV421 (clone J252D4), CCR7 FITC (clone G043H7), Ki67 PE (clone Ki-67), HLA-DR PE (clone L243), IL-21 APC (clone 3A3-N2), IFN-γ BV510 (clone B27), TNF-α FITC (clone Mab11), IL-17 BV421 (clone BL168) all from Biolegend and CXCL13 PE (clone 53610) from R&D Systems.

**TCRβ repertoire sequencing**

TCRβ rearrangements were amplified using amplicon rescue multiplex PCR and sequenced by next generation sequencing on an Illumina MiSeq platform (iRepertoire®, Huntsville, AL, USA). After initial filtering and mapping using iRepertoire® algorithms, multiple sequence copies of unique sequences were counted as a single sequence if not otherwise stated.

During the study, an in-house pipeline for TCRβ sequencing was established as well: cDNA was generated using the Applied Biosystems™ High-Capacity cDNA Reverse Transcription Kit per manufacturer's protocol. For semi-nested multiplex PCR primers from (1) were used: In a 1$^{st}$ PCR (15 cycles, annealing temperature 62 °C) a common linker sequence was introduced with the primers targeting the different TRBV segments (equimolar mix of 28 primers, final concentration of the mix 0.6 µM), the reverse primer targets the constant region of the TCRβ chain. In a 2$^{nd}$ PCR (50 cycles, annealing temperature 65 °C), with a forward primer targeting the linker sequence and a reverse primer binding to a more 5' part of the constant region (final concentration 0.5 µM each). The resulting product was purified using the QIAquick PCR Purification Kit (Quiagen) and sent for adapter ligation and amplicon sequencing to a commercial provider (GENEWIZ, Azenta Life Sciences, Amplicon-EZ with Illumina MiSeq 2x250 bp). Paired-end reads were assembled, quality filtered (mean Phred-Score >20) and deduplicated using the pRESTO workflow (2). Sequences occurring less than twice were excluded from further analysis.

**TCRβ repertoire analysis**

Resulting sequence data was analyzed using IMGT/HighV-QUEST, unproductive sequences were filtered out. Resulting files were further analyzed for V-, D-, and J-segment usage, CDR3-length, and clonality using ARGalaxy (3). TCR clonotypes were defined by identical V-, D- and J-gene segment usage as well as identical CDR3 nucleotide sequence. For calculation of clonal diversity resulting Change-O databases were analyzed using Alakazam (4). Standard settings were used for computation of the diversity scores (bootstrap n=200, ci=0.95). Immunarch was used for visualization of clonotype tracking and repertoire overlap (5). AIRR output tables from IMGT/HighV-QUEST (6) were adopted for use in Grouping of Lymphocyte Interactions by Paratope Hotspots version 2 (GLIPH2, (7)). Standard CD4 reference version 2.0 was used. Output data from GLIPH2 specificity groups with sequences from at least two patients was visualized as networks using the Cytoscape v3.9.1 software. Sequence logos were generated using the 'ggseqlogo' R vignette. Additionally, the frequencies of aa doublets at specific CDR positions were computed: 'GH' at the 2$^{nd}$ and 3$^{rd}$ positions in CDR1β (IMGT position 28 and 29) and 'SL' or 'SV' at the 8$^{th}$ and 7$^{th}$ positions from the end of the CDR3β-junction (including Cys-104 and Trp/Phe-118; IMGT position 111 and 112 for a junction length of 15 amino acids (aa) / CDR3β length of 13 aa). Sequences with a junction or CDR3β length less than 13 or 11 aa, respectively, were excluded from the analysis.

**10x Genomics Chromium single-cell RNA-seq**

Subsequent analysis on scRNA-seq output files was carried out using Seurat V4 for gene expression analysis (8) and scRepertoire version 1.7.2 for integrating VDJ sequencing data (9). Multiplexed samples were demultiplexed using cellsnp-lite and vireo and assigned to donors by combining this data with Hashtag Oligo UMIs in each donor (10). Quality control was done by calculating the median + 3*(median absolute deviation) from RNA counts, number of features and the percentage of mitochondrial reads per cell. Droplets with properties above these thresholds and below 2500/1000 RNA counts/feature counts were excluded from further analysis.

To focus the analysis on CD4$^{+}$ T cells each sample was normalized and clustered with the standard settings from Seurat (log-Normalization, 2000 most variable features) and clusters with expression of specific genes for B cells ("CD79A", "CD79B", "CD19", "MS4A1"), monocytes ("LYZ") and γδ T cells ("TRGC1", "TRGC2", "TRDC") were excluded from further analysis. TCR gene segment related features were removed from gene expression matrixes to avoid impact on clustering. The three samples were subsequently integrated using the data integration vignette of Seurat after normalization by the SCTransform function. Cell clusters were defined based on gene expression analysis using FindCluster with a resolution of 0.2. Differentially expressed genes were identified by the FindAllMarkers function of Seurat. GSEA analysis was done using the 'fgsea' R package with Reactome pathways. For creation of expression scores genes from Reactome pathways were used as input for AddModuleScore function from Seurat.

TCR data was integrated from the CellRanger output files using scRepertoire. Sequences from cells with more than one α and β chain each were excluded. Clonotypes were defined as having the same VDJC genes comprising the TCR and the same nucleotide sequence of the CDR3 region. Clonal networks and occupied repertoire space were analyzed using the built-in functions of scRepertoire. Clones (here same VJ-segment usage and same CDR3 aa sequence) were defined as being convergent when consisting of at least two unique CDR3 nucleotide sequences as proposed in (11). For velocity analyses, loom files with information on spliced and unspliced reads were computed from Cellranger output using velocyto and then further processed using scVelo in dynamic mode (12, 13).

**Adaption of published TCRβ data**

Publicly available TCRβ repertoire sequencing data sets were re-analyzed for this study.

Henderson *et al.* (14): Processed TCR repertoire data from FACS sorted $CD4^{+}CD25^{-}$ synovial fluid (SF) T cells of 11 JIA patients and two patients with LA were downloaded from www.adaptivebiotech.com/pub/Henderson-2015 and filtered for productive sequences. CDR1β and CDR3β amino acid sequences and TRBV/TRBJ assignments were used for further analysis.

Maschmeyer *et al.* (15): Published Cellranger output ‘filtered_contig.fasta’ files (10X genomics 5’ VDJ single cell sequencing, GEO: GSE160097) from FACS sorted SF $CD4^{+}CD45RO^{+}CD25^{-}$ of 7 JIA patients were processed with IMGT/HighV-Quest (6) using standard settings. Resulting AIRR-tables – filtered for productive TCRβ sequences – were used for further analysis.

Saguraki *et al.* (16): TCRβ bulk sequencing data from FACS sorted SF $CD4^{+}PD\text{-}1^{hi}CXCR5^{-}$ T cells of three RA patients was downloaded as raw reads from DDJB archive DRA011207. Raw read assembly and quality control was done using the pRESTO pipeline with standard settings (2). Sequences with at least two occurrences were used as input for IMGT/HighV-Quest and further analyzed as described above.

Fischer, Dirks *et al.* (17): TCRβ bulk sequencing data from FACS sorted SF $CD4^{+}PD\text{-}1^{hi}CXCR5^{-}$ T cells of four JIA patients processed in our laboratory in a previous study (Genbank KEXF01000000) were further analyzed as described above.

TCRs with known antigen specificities/VDJdb database:

To elucidate CDR3β motifs implicated in the binding of viral antigens, a work-flow similar to that described in (18) was employed. Specifically, a total of 55,471 TCR sequences possessing documented specificities for EBV, CMV or Influenza A were retrieved from the VDJdb database as of January 26, 2023 (19) and utilized as input data for GLIPH2 analysis. The criteria applied for pattern settings involved the selection of sequences with unique CDR3 counts greater than 4, Fisher scores below 0.05, as well as vb and length scores lower than 0.05, collectively forming what is hereafter referred to as the 'viral pattern'. Subsequently, cells identified within the ARLA scRNA-seq dataset of this study, exhibiting the presence of any of these defined viral patterns, were categorized into the 'viral motives' group.

To investigate the presence of ARLA-associated TCRβ motives among TCRs exhibiting known specificities, a search was conducted within the VDJdb database (19). This search targeted TCRβ sequences featuring MHCII restrictions and encompassing over ten TCRs aimed at antigens within a particular organism.

SUPPLEMENTAL TABLES

| Patient ID | Sex | Affected joints | Borrelia serology (IB) IgM | IgG | ANA | HLA-DRB1 | HLA-B27 | Antibiotic Treatment |
|---|---|---|---|---|---|---|---|---|
| **ARLA01** | male | knee | negative | p100 (+), VlsE (+), p58 (+), p41 (+), p39 (+), OspA (-), OspC (+), p18 (+) | 1:160 | 11:01 11:01 | negative | ceftotaxime (2 weeks), ceftotaxime (2 weeks) |
| **ARLA02** | female | knee | negative | p100 (+), VlsE (+), p58 (+), p41 (+), p39 (-), OspA (-), OspC (+), p18 (+) | 1:80 | 11:01 11:02 | negative | amoxicilline (4 weeks), ceftriaxone (2 weeks) |
| **ARLA03** | male | knee | negative | p100 (+), VlsE (+), p58 (+), p41 (+), p39 (-), OspA (-), OspC (+), p18 (+) | <1:80 | 12:01 13:02 | negative | amoxicilline (3 weeks), doxycycline (4weeks) |
| **ARLA04** | female | knee | negative | p100 (+), VlsE (+), p58 (+), p41 (+), p39 (+), OspA (-), OspC (+), p18 (+) | 1:2560 | 01:01 04:01 | negative | cefuroxime (2 weeks), doxycycline (4 weeks) |
| **ARLA05** | male | knee | p41 (+), Osp17 (+), VlsE (+) | VlsE (+), p41 (+), p41 (+), p39 (+), p21 (+), Osp17 (+), p18 (+), p14 (+) | 1:80 | 11:04 15:04 | negative | doxycycline (8 weeks), ceftriaxone (2 weeks) |
| **ARLA06** | male | knee | p41 (+), p30 (+), p25 (+) | VlsE (+), p83 (+), p41 (+), p39 (+), p34 (+) | <1:80 | 11:02 07:01 | negative | doxycycline (4 weeks), ceftriaxone (2 weeks) |
| **ARLA07** | male | knee | OspC (+) | p100 (+), VlsE (+), p58 (+), p41 (+), p39 (+), OspA (-), OspC (-), p18 (+) | <1:80 | 07:01 13:01 | negative | ceftriaxone (2 weeks) ceftriaxone (2 weeks) |
| **ARLA08** | male | knee | negative | p83 (+), p58 (+), p21 (+), p43 (+), p39 (+), p30 (+), p25 (+) | 1:160 | 04:04 08:04 | negative | doxycycline (3 weeks), ceftriaxone (3 weeks) |
| **ARLA09** | female | knee | OspC (+) | p100 (+), VlsE (+), p58 (+), p41 (+), p39 (+), OspA (-), OspC (+), p18 (+) | <1:80 | 07:01 08:01 | negative | cefotaxime (4 weeks), amoxicilline (4 weeks) |
| **ARLA10** | female | knee | negative | p100 (+), VlsE (+), p58 (+), p41 (+), p39 (+), OspA (-), OspC (+), p18 (+) | 1:80 | 04:01 07:01 | positive | ceftriaxone (2 weeks), amoxicilline (4 weeks |
| **ARLA11** | male | knees | p41 (+), OspC (+) | p100 (+), VlsE (+), p58 (+), p41 (+), p39 (+), OspA (-), OspC (+), p18 (+) | <1:80 | 01:01 07:01 | positive | doxycycline (4 weeks), ceftriaxone (2 weeks) |
| **ARLA12** | male | knee | negative | p100 (+), VlsE (+), p58 (+), p41 (+), p39 (+), OspA (-), OspC (+), p18 (+) | <1:80 | 11:04 13:223 | negative | doxycycline (4 weeks), ceftriaxone (2 weeks) |
| **ARLA13** | female | knee | negative | p100 (+), VlsE (+), p58 (+), p41 (+), p39 (+), OspA (+), OspC (+), p18 (+) | 1:120 | 15:01 15:01 | negative | amoxicilline (4 weeks), doxycycline (4weeks) |

**Supplemental Table 1 – Clinical and demographical characterization of ARLA patients**

| *HLA-DRB1* allele | North America | | | | | Germany | | |
|---|---|---|---|---|---|---|---|---|
| | Responsive (n=100) | | Refractory (n=142) | | Control (n=3,798) | Refractory (n=26) | | Control (n=3,456,066) |
| | number | % | number | % | % | number | % | % |
| **01:01** | 4 | 4.0 | 16 | 11.3 | 9.1 | 2 | 7.7 | 9.7 |
| **04:01** | 6 | 6.0 | 18 | 12.7 | 10.3 | 2 | 7.7 | 8.2 |
| **15:01** | 9 | 9.0 | 20 | 14.1 | 16.3 | 2 | 7.7 | 13.3 |
| | 19 | **19.0** | 54 | **38.1** | **35.7** | 6 | **23.1** [1] | **31.2** |
| **08:01** | 6 | 6.0 | 1 | 0.7 | 2.2 | 1 | 3.9 | 2.9 |
| **08:04** | 0 | 0.0 | 0 | 0.0 | 0.0 | 1 | 3.9 | 0.2 |
| **11:01** | 9 | 9.0 | 4 | 2.8 | 5.6 | 3 | 11.5 | 7.8 |
| **11:02** | 0 | 0.0 | 1 | 0.7 | 0.0 | 2 | 7.7 | 0.2 |
| **11:04** | 7 | 7.0 | 2 | 1.4 | 2.7 | 2 | 7.7 | 2.9 |
| **13:01** | 4 | 4.0 | 6 | 4.2 | 5.6 | 1 | 3.9 | 7.6 |
| **13:02** | 7 | 7.0 | 3 | 2.1 | 4.1 | 1 | 3.9 | 4.2 |
| | 33 | **33.0** | 17 | **11.9** | **20.2** | 10 | **42.5** [2] | **25.8** |

**Supplemental Table 2 – Distribution of HLA-DRB1 allele frequencies**

The distribution of selected HLA-DRB1 alleles is shown within the cohort of patients with antibiotic-refractory Lyme Arthritis (ARLA) analyzed in this study (Germany, Refractory) as well as data obtained from a previous study involving Caucasian patients from North America with antibiotic-responsive and refractory Lyme Arthritis (20). Data from German control individuals were retrieved from the population of ‘Germany DKMS – German donors’ from the ‘Allele frequency net database’ (21). Data from North American control individuals were retrieved from European-American bone marrow donors (20, 22). The upper part of the table reports the known ‘risk alleles’ identified in the North American cohort, while the lower section summarizes the known ‘protective alleles’. Comparative analysis of cumulative frequencies for each allele group was conducted between both antibiotic-refractory patient cohorts using Fisher`s exact test ([1] $p=0.18$; [2] $p<0.01$).

| | Laboratory assessments | | | | | |
|---|---|---|---|---|---|---|
| **Patient ID** | **HLA-DRB1** | **Distribution PD-1$^{hi}$HLA-DR$^{+}$ (FACS)** | **Follow-up distribution PD-1$^{hi}$HLA-DR$^{+}$ (FACS)** | **TRVB sequencing in PD-1$^{hi}$HLA-DR$^{+}$** | **Follow-up TRVB sequencing in PD-1$^{hi}$HLA-DR$^{+}$** | **scRNA-Seq CD4$^{+}$ cells** |
| **ARLA01** | 11:01<br>11:01 | yes | yes | yes | yes | yes |
| **ARLA02** | 11:01<br>11:02 | yes | yes | yes | yes | yes |
| **ARLA03** | 12:01<br>13:02 | yes | no | yes | no | no |
| **ARLA04** | 01:01<br>04:01 | yes | no | yes | no | no |
| **ARLA05** | 11:04<br>15:04 | yes | yes | yes | yes | yes |
| **ARLA06** | 11:02<br>07:01 | yes | no | yes | no | no |
| **ARLA07** | 07:01<br>13:01 | yes | no | yes | no | no |
| **ARLA08** | 04:04<br>08:04 | yes | no | yes | no | no |
| **ARLA09** | 07:01<br>08:01 | yes | no | yes | no | no |
| **ARLA10** | 04:01<br>07:01 | yes | yes | no | no | no |
| **ARLA11** | 01:01<br>07:01 | yes | no | yes | no | no |
| **ARLA12** | 11:04<br>13:223 | yes | no | yes | no | no |
| **ARLA13** | 15:01<br>15:01 | yes | yes | yes | no | no |

**Supplemental Table 3 – Experimental analyses conducted in ARLA patients**

**Supplemental Table 4, 5 and 6** are provided in 'supp_tables.xlsx'

SUPPLEMENTAL FIGURES

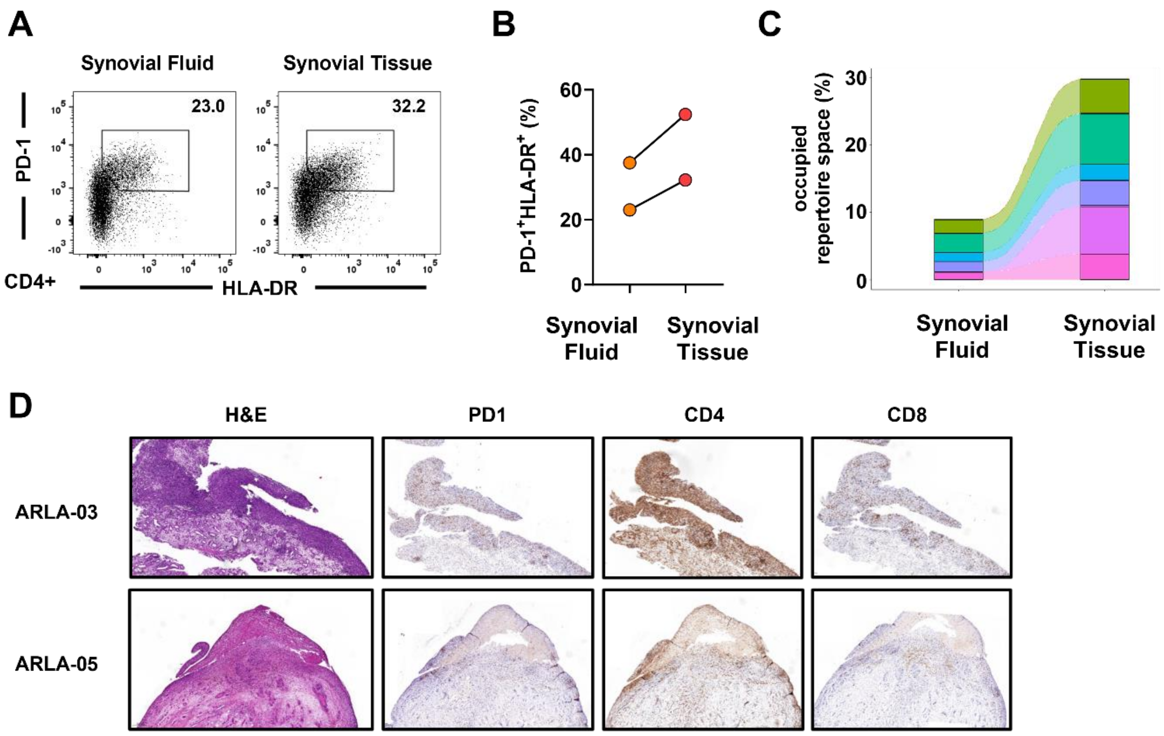

**Supplemental Figure 1 – Distribution of PD-1$^{hi}$HLA-DR$^{+}$ CD4$^{+}$ T cells and TCR clonal overlap between synovial fluid and synovial tissue**

**(A)** Representative dot plots and **(B)** frequencies of PD-1$^{hi}$HLA-DR$^{+}$ CD4$^{+}$ T cells in matched synovial fluid and synovial tissue of two ARLA patients. **(C)** Proportion of occupied repertoire space (reads per total reads) of overlapping clonotypes (same V gene and same CDR3 amino acid, determined by ARGalaxy) in PD-1$^{hi}$HLA-DR$^{+}$CD4$^{+}$ T cells from synovial fluid and synovial tissue of patient ARLA05. **(D)** Representative histological and immunohistological analysis of synovial tissue sections from two ARLA patients.

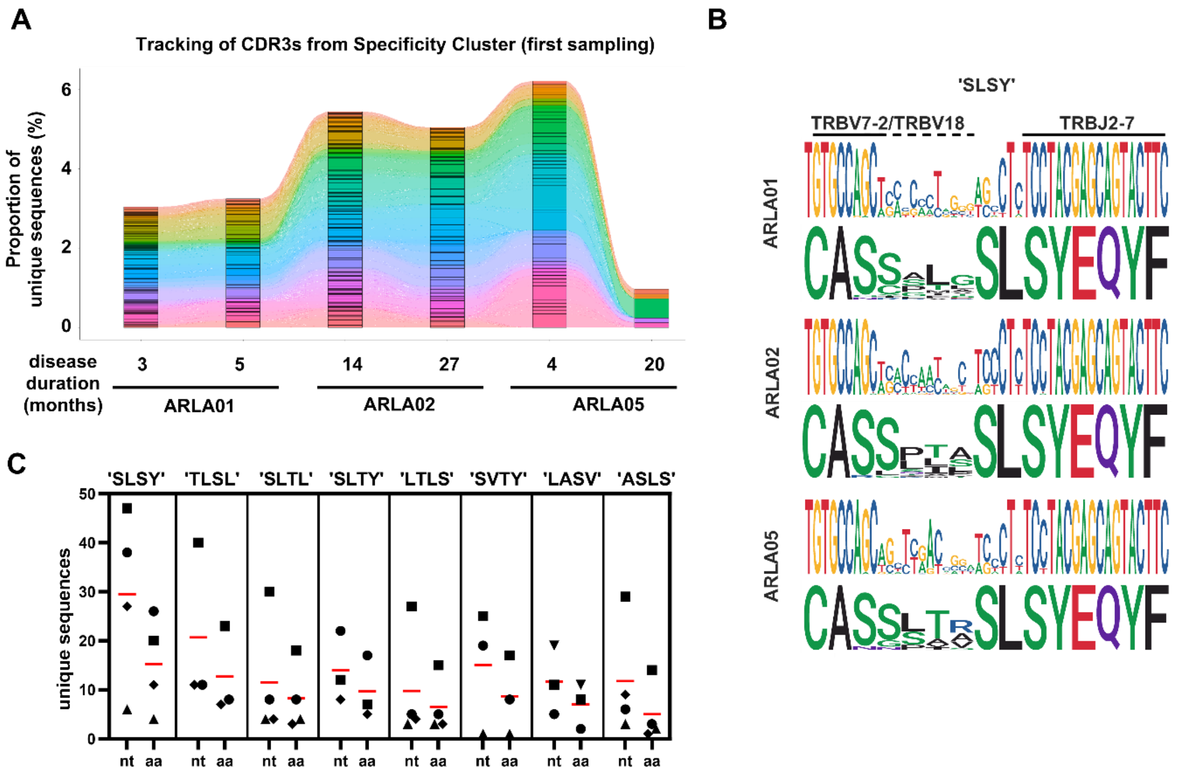

**Supplemental Figure 2 – The TCR CDR3β motive enriched in synovial fluid PD-1$^{hi}$HLA-DR$^{+}$CD4$^{+}$ T cells in ARLA patients is non-germline encoded**

**(A)** Tracking of occupied repertoire space using CDR3β amino acid (aa) sequences derived from the 'specificity cluster' (Figure 2B) at various time points in three patients with available follow-up samples. Each color corresponds to a unique CDR3β aa sequence. **(B)** Sequence plots of the CDR3β joining region depict nucleotide (nt) sequences (each in the upper row) and their corresponding aa sequences (each in the lower row) for motifs containing 'SLSY' in the CDR3β, organized separately for each patient. **(C)** The absolute counts of unique nt or aa sequences resulting in CDR3βs with specified aa motifs (motifs selected from the 'specificity cluster,' filtered for an overall count of unique CDR3β (aa) ≥ 20), each symbol represents one patient.

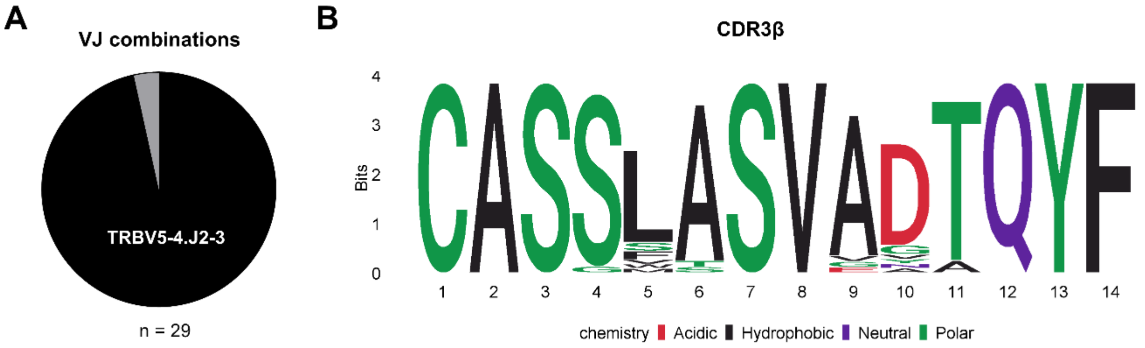

**Supplemental Figure 3 – ARLA-associated TCRβ sequences from patient ARLA06 reveal a different TRBV-TRBJ gene usage bias**

**(A)** The percentage of TBRV5-4.TBRJ2-3 pairing within TCRβ sequences featuring the ARLA-associated CDR3β motif ('SL'/'SV' at IMGT-position 111/112) obtained from patient ARLA06. **(B)** Sequence logos displaying the amino acid sequences of CDR3β derived from the aforementioned sequences in (A).

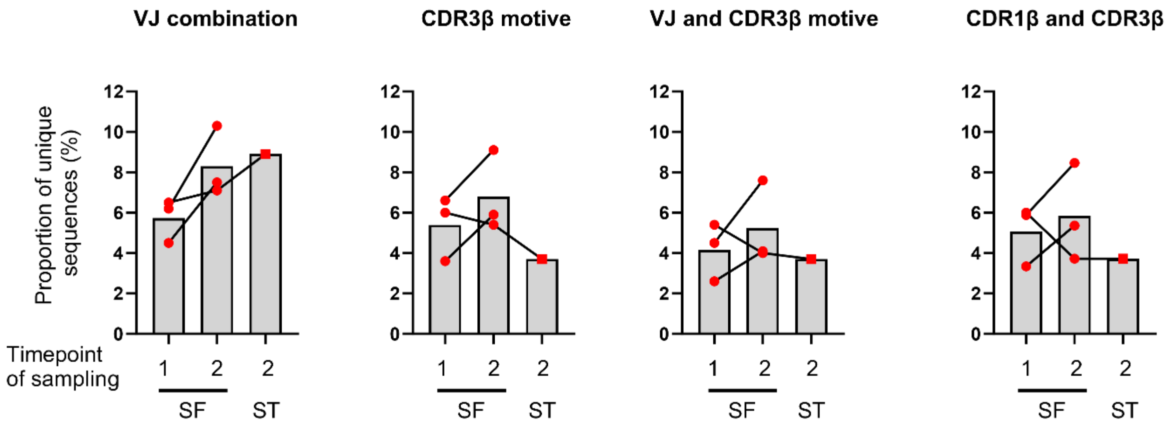

**Supplemental Figure 4 – T cell clones bearing ARLA-associated TCRβ motives persist during disease course and are present in synovial tissue**

Frequencies of indicated TCRβ motives in synovial fluid PD-1$^{hi}$HLA-DR$^{+}$CD4$^{+}$ T cells from ARLA patients with available follow-up samples (n=3) and available synovial tissue samples (n=1) determined by bulk sequencing.

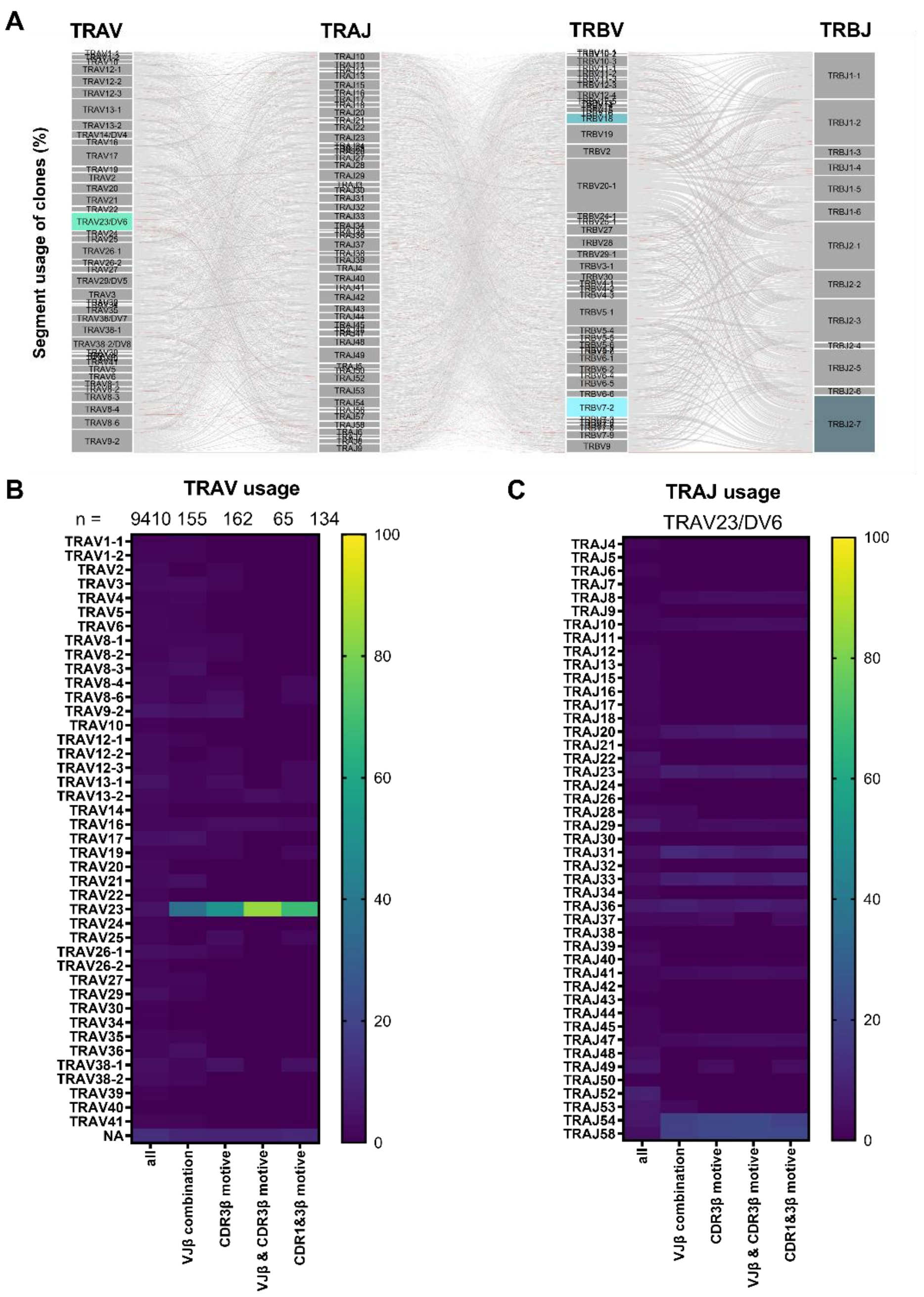

**Supplemental Figure 5 - Paired TCRα/β-sequencing reveals strong bias in TRAV gene segment usage in TCRs with ARLA-associated TCRβ motives**

**(A)** Alluvial plot illustrating TRAV-TRAJ-TRBV-TRBJ combinations (determined through paired TCRα/β-sequencing of synovial fluid CD4$^+$ T cells from three ARLA patients) in unique clones devoid of motives from the ‘specificity cluster’ in the CDR3β. Clones bearing the CDR3β motive (‘SL’/’SV’ at IMGT-position 111/112 in CDR3β) are highlighted in coral. **(B)** Heat map displaying the TRAV usage in unique clones obtained from paired TCRα/β-sequencing of SF CD4$^+$ T cells from three ARLA patients, categorized into either all clones or specific subgroups based on TCRβ properties. The number of clones in each group is displayed at the top. **(C)** Heat map representing TRAJ usage in the clones from (B) that utilize the TRAV23/DV6 combination, delineated across the same subgroups.

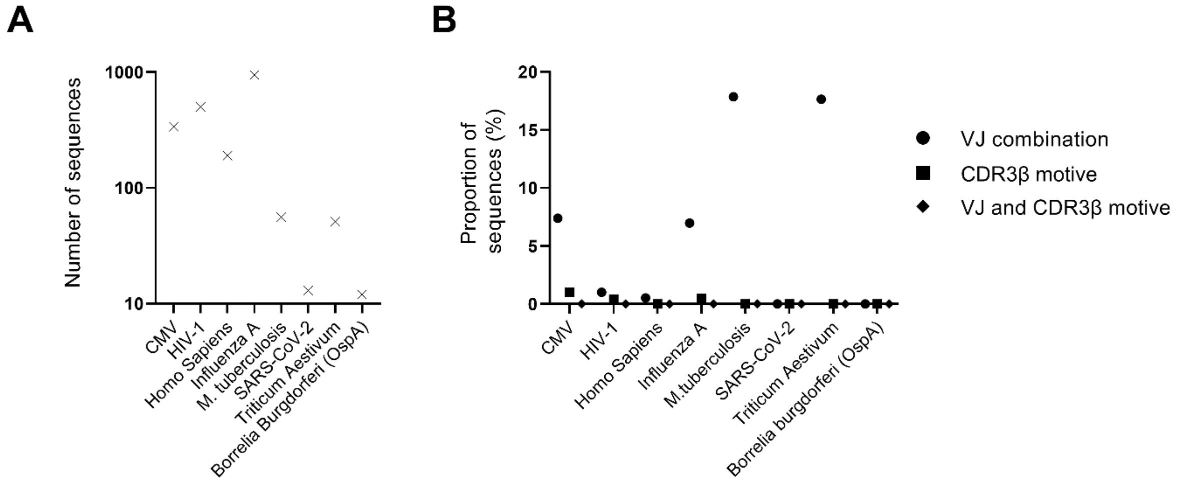

**Supplemental Figure 6 – Search of ARLA-associated TCRβ motives in public TCR databases**

**(A)** Number of TCRβ sequences with known antigen specificity from the vdjdb (filtered for antigens with n > 10 sequences and MHCII context). TCRβ sequences specific for *Borrelia burgdorferi* ss outer surface protein A (OspA) are retrieved from (23). **(B)** Frequencies of indicated ARLA-associated TCRβ motives in TCRβ sequences from (A).

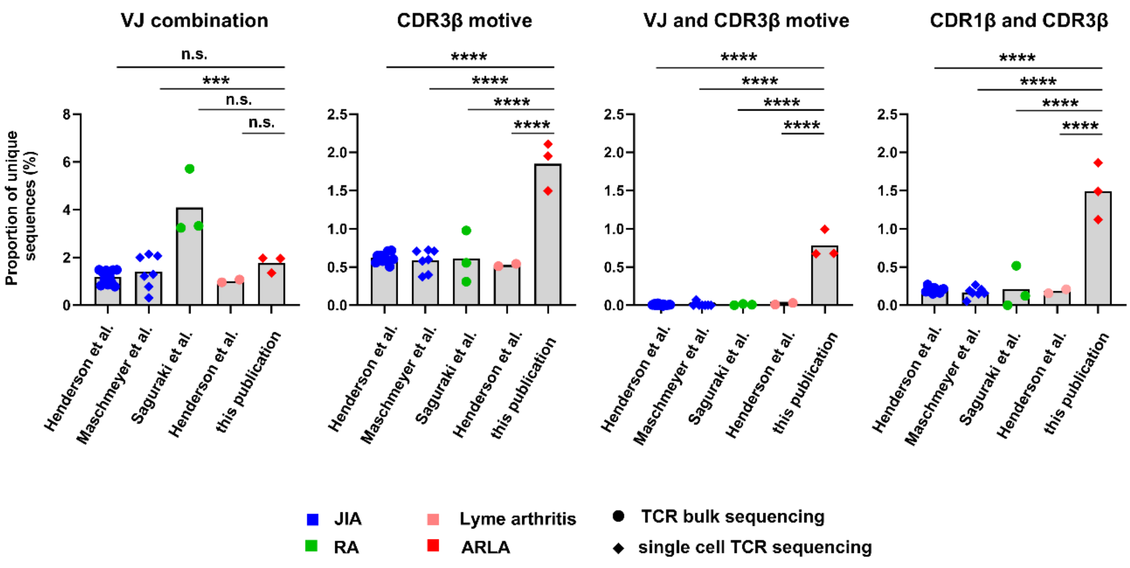

**Supplemental Figure 7 – ARLA-associated TCRβ motives are not present in synovial fluid CD4$^+$ T cell clones in other forms of chronic arthritis**

Frequencies of indicated TCRβ motives in synovial fluid (SF) CD4$^+$ T conventional cells from patients with juvenile idiopathic arthritis (JIA), rheumatoid arthritis (RA), Lyme arthritis (LA) and ARLA (Henderson et al.: JIA, n=11, FACS sorted CD4$^+$CD25$^-$CD127$^{+/-}$ SF T cells, bulk sequencing (14); Maschmeyer et al.: JIA, n=7, FACS sorted CD4$^+$CD45RO$^+$CD25$^-$CD127$^{+/-}$ SF T cells, single cell RNA + VDJ sequencing (15); Saguraki et al.: RA, n=3, FACS sorted CD4$^+$PD-1$^{high}$CXCR5$^-$ SF T cells, bulk sequencing (14); Henderson et al.: LA, n=2, FACS sorted CD4$^+$CD25$^-$CD127$^{+/-}$ SF T cells, bulk sequencing (14); this publication: ARLA, n=3, SF CD4$^+$ T cells, single cell RNA + VDJ sequencing). One-way ANOVA and multiple comparisons (to ‘ARLA’) corrected with Dunnett’s formula; ***: p<0.001, ****: p<0.0001.

| Patient ID | TCRβ motive (%) | HLA-DRB1 alleles | HLA-DRB1 amino acid position 9 | 11 | 13 | 26 | 28 | 30 | 37 | 47 | 57 | 58 | 61 | 66 | 67 | 69 | 70 | 71 | 72 | 73 | 74 | 77 | 78 | 81 | 82 | 85 | 86 | 89 | 90 |
|---|---|---|---|---|---|---|---|---|---|---|---|---|---|---|---|---|---|---|---|---|---|---|---|---|---|---|---|---|---|
| | | | Peptide binding pocket 9 | 6 | 4 | 4 | 4+7 | 6 | 9 | 7 | 9 | | 7 | | 7 | | 4 | 4 | | | 4 | | 4 | | | 1 | 1 | 1 | 1 |
| 06 | 6.7 | 11:02 | E | S | S | F | D | Y | Y | F | D | E | W | D | I | E | D | E | R | A | A | T | Y | H | N | V | V | F | T |
| | | 07:01 | E | G | Y | F | E | L | F | Y | V | A | W | D | I | E | D | R | R | G | Q | T | V | H | N | V | G | F | T |
| 05 | 6.0 | 11:04 | E | S | S | F | D | Y | Y | F | D | E | W | D | F | E | D | R | R | A | A | T | Y | H | N | V | V | F | T |
| | | 15:04 | W | P | R | F | D | Y | S | F | D | A | W | D | F | E | Q | A | R | A | A | T | Y | H | N | V | V | F | T |
| 02 | 5.9 | 11:01 | E | S | S | F | D | Y | Y | F | D | E | W | D | F | E | D | R | R | A | A | T | Y | H | N | V | G | F | T |
| | | 11:02 | E | S | S | F | D | Y | Y | F | D | E | W | D | I | E | D | E | R | A | A | T | Y | H | N | V | V | F | T |
| 03 | 3.5 | 13:02 | E | S | S | F | D | Y | N | F | D | A | W | D | I | E | D | E | R | A | A | T | Y | H | N | V | G | F | T |
| | | 12:01 | E | S | G | L | E | H | L | F | V | A | W | D | I | E | D | R | R | A | A | T | Y | H | N | A | V | F | T |
| 01 | 3.4 | 11:01 | E | S | S | F | D | Y | Y | F | D | E | W | D | F | E | D | R | R | A | A | T | Y | H | N | V | G | F | T |
| | | 11:01 | E | S | S | F | D | Y | Y | F | D | E | W | D | F | E | D | R | R | A | A | T | Y | H | N | V | G | F | T |
| 12 | 2.8 | 11:04 | E | S | S | F | D | Y | Y | F | D | E | W | D | F | E | D | R | R | A | A | T | Y | H | N | V | V | F | T |
| | | 13:223 | E | S | S | F | D | Y | N | F | D | A | W | D | I | E | D | E | R | A | A | T | Y | H | N | V | V | F | T |
| 07 | 0.5 | 13:01 | E | S | S | F | D | Y | N | F | D | A | W | D | I | E | D | E | R | A | A | T | Y | H | N | V | V | F | T |
| | | 07:01 | W | G | Y | F | E | L | F | Y | V | A | W | D | I | E | D | R | R | G | Q | T | V | H | N | V | G | F | T |
| 11 | 0.4 | 01:01 | W | L | F | L | E | C | S | Y | D | A | W | D | L | E | Q | R | R | A | A | T | Y | H | N | V | G | F | T |
| | | 07:01 | W | G | Y | F | E | L | F | Y | V | A | W | D | I | E | D | R | R | G | Q | T | V | H | N | V | G | F | T |
| 08 | 0.2 | 04:04 | E | V | H | F | D | Y | Y | Y | D | A | W | D | L | E | Q | R | R | A | A | T | Y | H | N | V | V | F | T |
| | | 08:04 | E | S | G | F | D | Y | Y | Y | D | A | W | D | F | E | D | R | R | A | L | T | Y | H | N | V | V | F | T |
| 09 | 0.2 | 07:01 | W | G | Y | F | E | L | F | Y | V | A | W | D | I | E | D | R | R | G | Q | T | V | H | N | V | G | F | T |
| | | 08:01 | E | S | G | F | D | Y | Y | Y | S | A | W | D | F | E | D | R | R | A | L | T | Y | H | N | V | G | F | T |
| 13 | 0.2 | 15:01 | W | P | R | F | D | Y | S | F | D | A | W | D | I | E | Q | A | R | A | A | T | Y | H | N | V | V | F | T |
| | | 15:01 | W | P | R | F | D | Y | S | F | D | A | W | D | I | E | Q | A | R | A | A | T | Y | H | N | V | V | F | T |
| 04 | 0.0 | 01:01 | W | L | F | L | E | C | S | Y | D | A | W | D | L | E | Q | R | R | A | A | T | Y | H | N | V | G | F | T |
| | | 04:01 | E | V | H | F | D | Y | Y | Y | D | A | W | D | L | E | Q | K | R | A | A | T | Y | H | N | V | G | F | T |
| 10 | na | 04:01 | E | V | H | F | D | Y | Y | Y | D | A | W | D | L | E | Q | K | R | A | A | T | Y | H | N | V | G | F | T |
| | | 07:01 | W | G | Y | F | E | L | F | Y | V | A | W | D | I | E | D | R | R | G | Q | T | V | H | N | V | G | F | T |

**Supplemental Figure 8 – Association of ARLA-associated TCRβ motives and HLA-DRB1 amino acid sequence**

ARLA patients are ranked based on the frequency of the TCR CDR1β+CDR3β motive. The positions of important amino acid (aa) residues corresponding to the HLA-DRB1 alleles are provided. Residues participating in the formation of a particular antigen binding pocket are indicate by the specific number of the pocket. Aa are represented by letters and color-coded. (na, not assessed); (24).

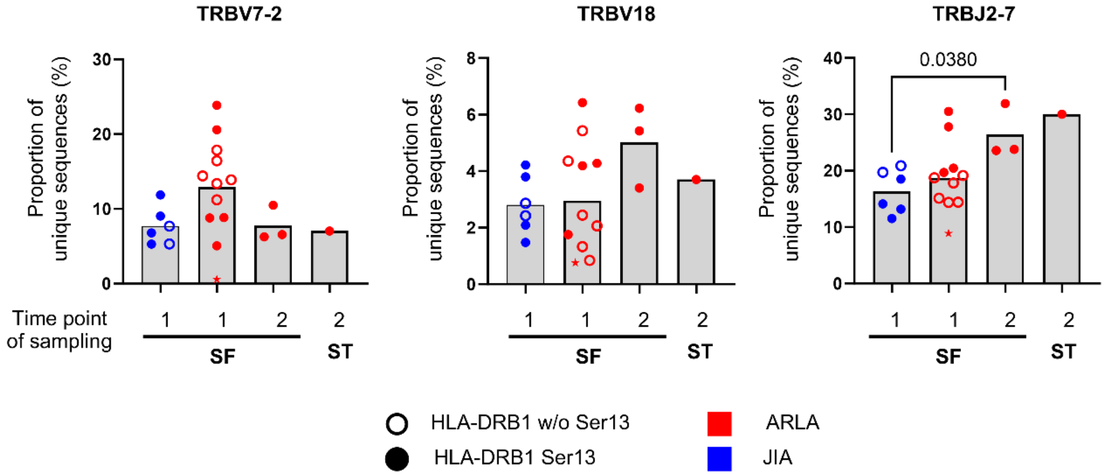

**Supplemental Figure 9 – Distribution of TCRβ VJ-segments**

Frequencies of usage of indicated TRBV and TRBJ gene segments in TCRβ sequences of synovial fluid (SF) and synovial tissue (ST) PD-1$^{hi}$HLA-DR$^{+}$CD4$^{+}$ T cells from children with juvenile idiopathic arthritis (JIA) and ARLA determined by bulk sequencing. Patients with Serine at position 13 (Ser13) of HLA-DRB1 on at least one allele are depicted with filled circles; ARLA06 (with presence of the CDRβ motive but distinct TRBV5-4.J2-3 pairing) is highlighted as red asterisk.

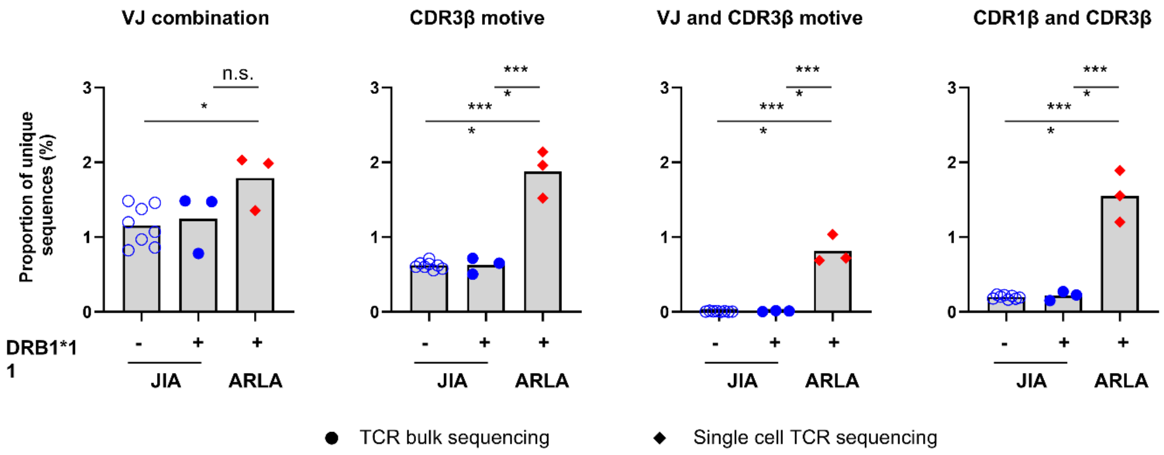

**Supplemental Figure 10 - Surrogate markers for ARLA-associated TCRs are disease specific and HLA-DRB1 restricted**

Frequencies of indicated TCRβ motives of synovial fluid (SF) $CD4^+$ T conventional cells from children with juvenile idiopathic arthritis (JIA) (n=11, FACS sorted $CD4^+CD25^-CD127^{+/-}$ SF T cells, bulk sequencing; this study and (17)) and ARLA (n=3, SF $CD4^+$ T cells, single cell RNA + VDJ sequencing, Treg cluster excluded for analysis). Patients with HLA-DRB1*11 genotype on at least one allele are depicted with filled symbols.

A

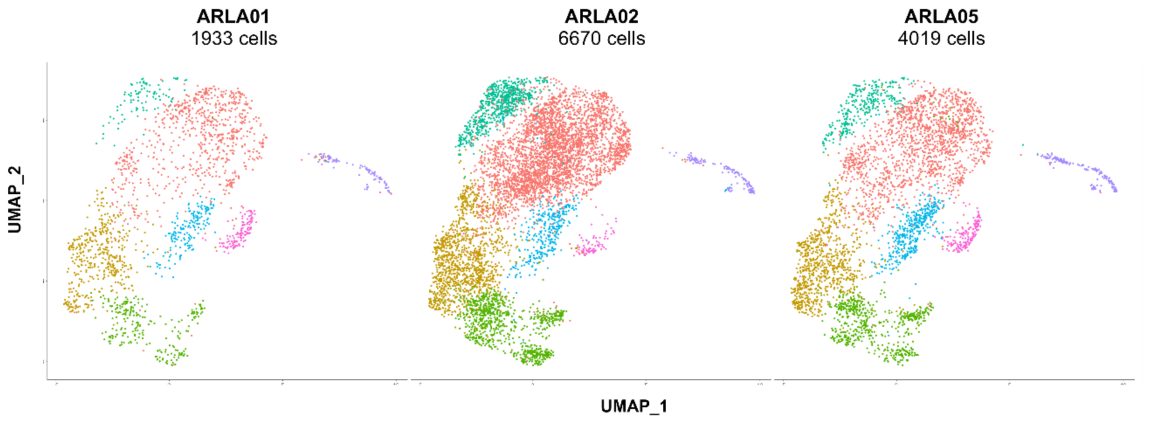

B

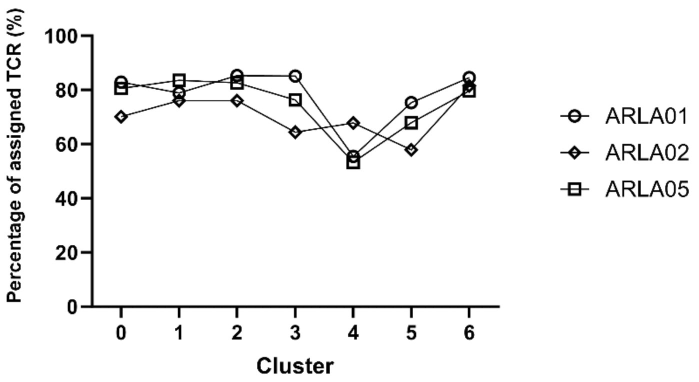

**Supplemental Figure 11 – Transcriptomic clusters and percentage of TCR assignment are equally distributed between patients**

**(A)** Individual UMAP representations, as depicted in Figure 5A, displaying transcriptome-based clustering separated for each patient **(B)** Percentage of cells within each cluster of a patient, wherein TCR sequences have been assigned following quality control measures.

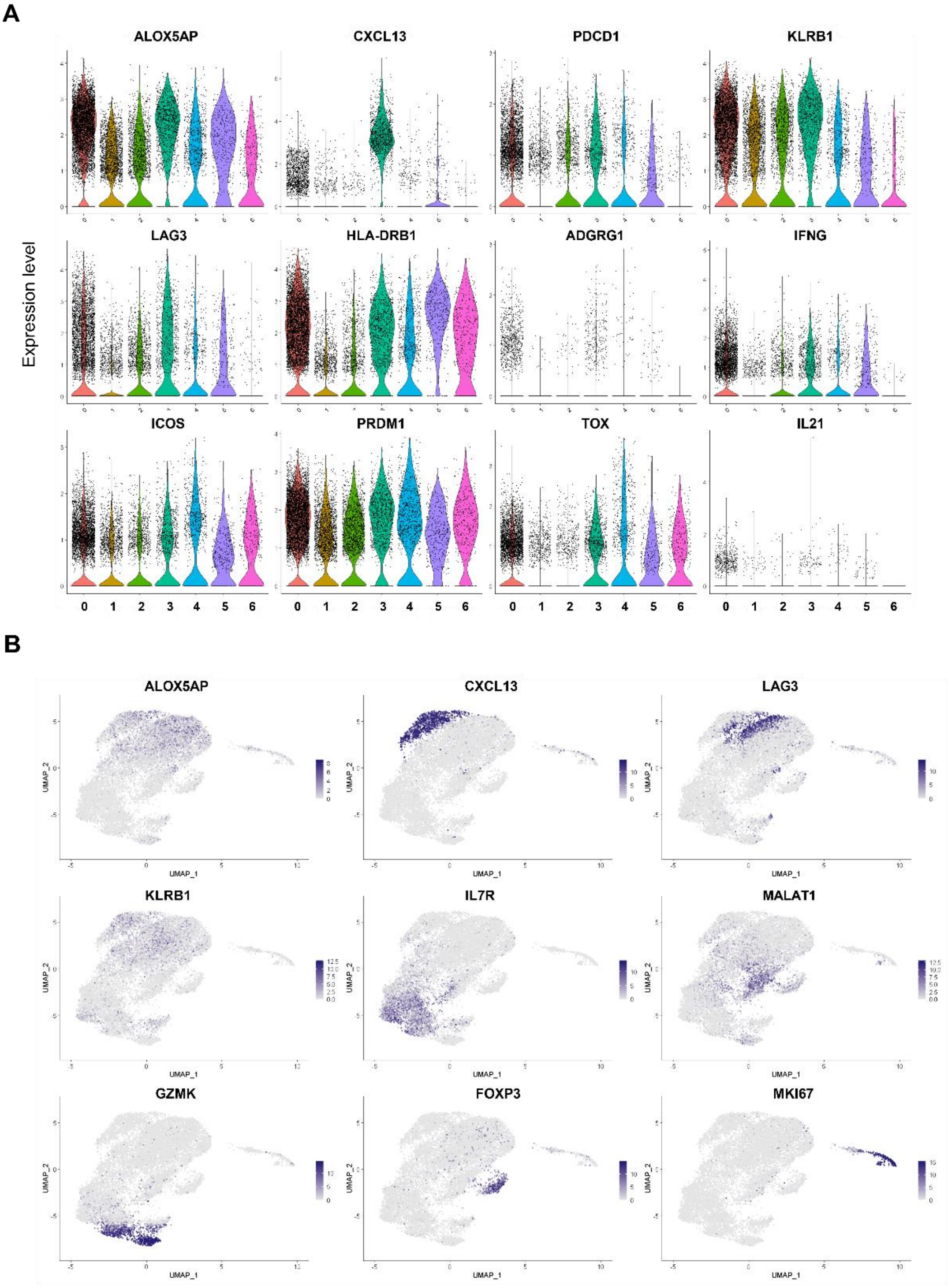

**Supplemental Figure 12 – Differential expression of marker genes associated in synovial fluid CD4⁺ T cell clusters**

**(A)** Violin plots showing the expression of indicated marker genes associated with peripheral T helper cells across the clusters. **(B)** UMAP representation with expression of selected marker genes characteristic for the respective cluster.

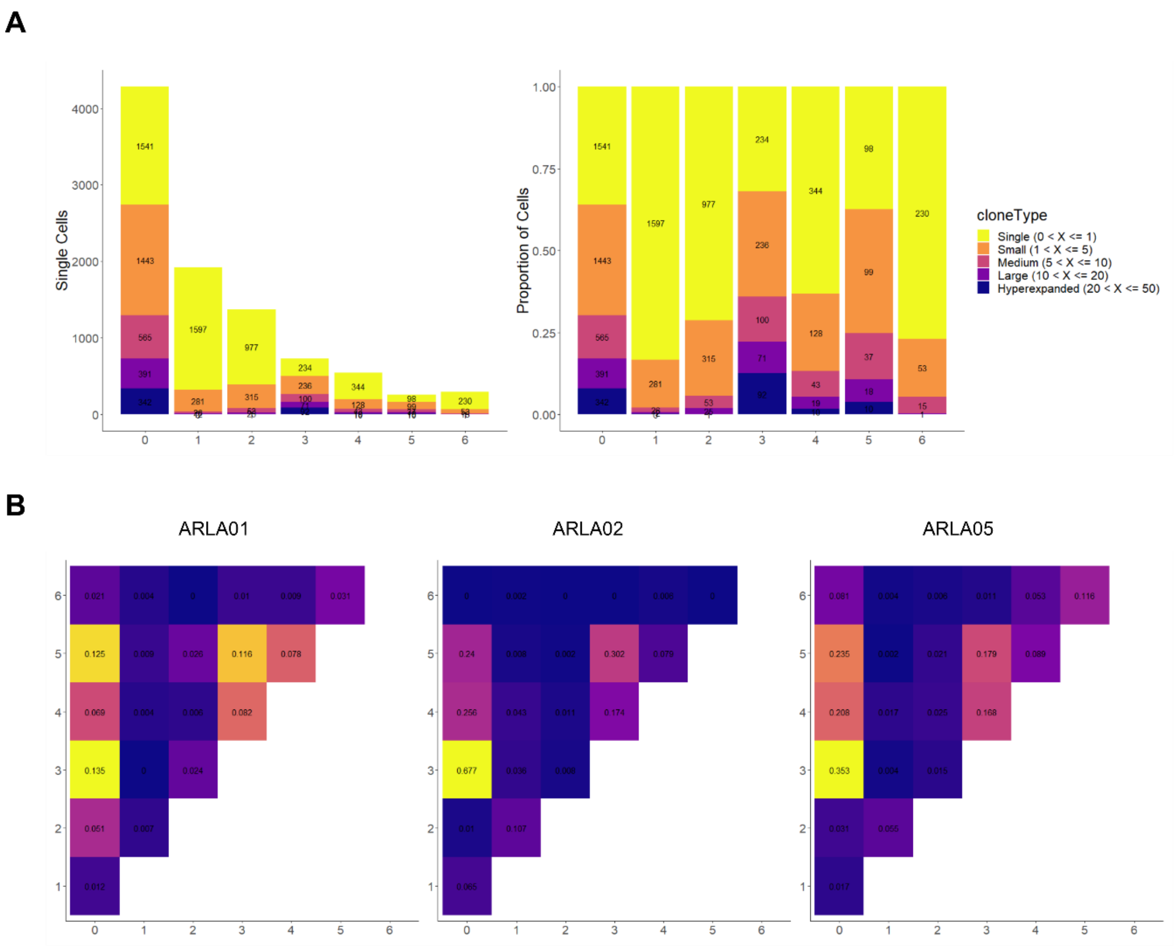

**Supplemental Figure 13 – Peripheral T helper cells exhibit clonal expansion in ARLA and patterns of clonal connectivity show similarity across different patients**

**(A)** Bar plot showing absolute and relative frequencies of cells with varying clone sizes within each cluster. **(B)** Morisita overlap index as a surrogate for clonal connectivity between clusters across the three patients.

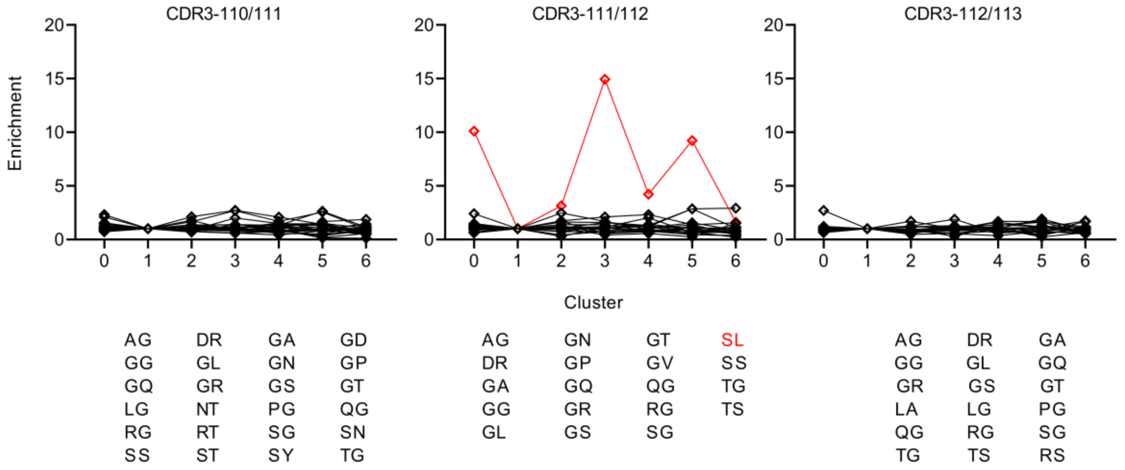

**Supplemental Figure 14 – Amino acid 'SL' doublet is enriched in peripheral T helper cell clusters compared to resting T cell clusters**

Unbiased visualization of fold enrichment of amino acid doublet frequencies at indicated CDR3β positions compared to cluster 1 ('resting' T cells). Doublets were filtered for frequencies > 1% of all doublets at the respective positions. The 'SL' doublet is highlighted in red.

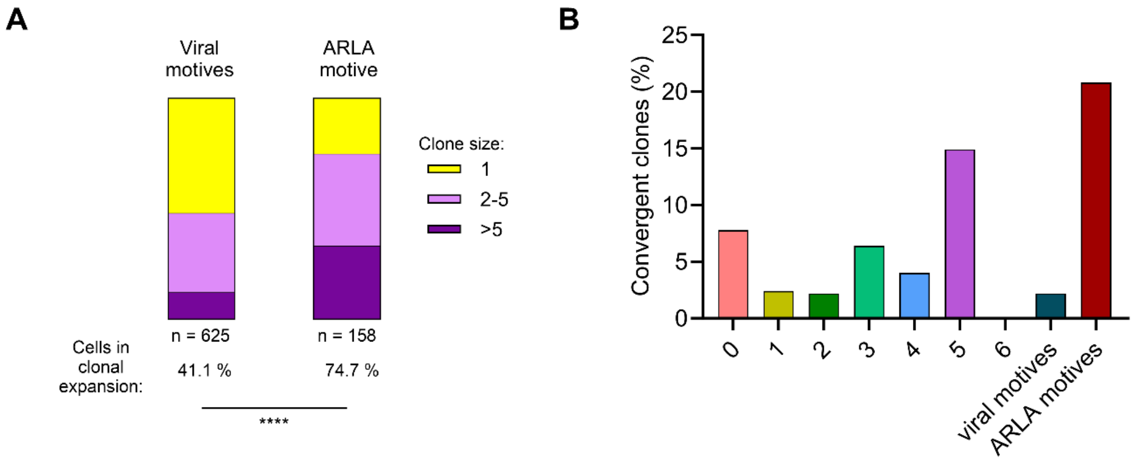

**Supplemental Figure 15 – Synovial fluid T cells bearing ARLA-associated TCRβ motives display signs of antigen-driven activation**

(A) Percentage of cells exhibiting various degrees of clonal expansion within the group of cells with 'viral' motives or the ARLA motive. Significance for cells in clonal expansion or not was calculated by Fisher's exact test between the two groups; ****: $p<0.0001$. (B) Frequency of convergent clones ($n \geq 2$ CDR3β nt sequences per clone) among all expanded clones (clone is defined as $n \geq 2$ cells with identical CDR3β aa and TRBV-TRBJ annotation) within respective clusters or groups of cells; TCR convergence according to (11).

SUPPLEMENTAL REFERENCES

1. Wahl I, Hoffmann S, Hundsdorfer R, Puchan J, Hoffman SL, Kremsner PG, et al. An efficient single-cell based method for linking human T cell phenotype to T cell receptor sequence and specificity. *Eur J Immunol.* 2022;52(2):237-46.
2. Vander Heiden JA, Yaari G, Uduman M, Stern JN, O'Connor KC, Hafler DA, et al. pRESTO: a toolkit for processing high-throughput sequencing raw reads of lymphocyte receptor repertoires. *Bioinformatics.* 2014;30(13):1930-2.
3. H IJ, van Schouwenburg PA, van Zessen D, Pico-Knijnenburg I, Stubbs AP, and van der Burg M. Antigen Receptor Galaxy: A User-Friendly, Web-Based Tool for Analysis and Visualization of T and B Cell Receptor Repertoire Data. *J Immunol.* 2017;198(10):4156-65.
4. Gupta NT, Vander Heiden JA, Uduman M, Gadala-Maria D, Yaari G, and Kleinstein SH. Change-O: a toolkit for analyzing large-scale B cell immunoglobulin repertoire sequencing data. *Bioinformatics.* 2015;31(20):3356-8.
5. Nazarov V, Tsvetkov V, Fiadziushchanka S, Rumynskiy E, Popov A, Balashov I, et al. https://immunarch.com/, https://github.com/immunomind/immunarch; 2023.
6. Alamyar E, Duroux P, Lefranc MP, and Giudicelli V. IMGT((R)) tools for the nucleotide analysis of immunoglobulin (IG) and T cell receptor (TR) V-(D)-J repertoires, polymorphisms, and IG mutations: IMGT/V-QUEST and IMGT/HighV-QUEST for NGS. *Methods Mol Biol.* 2012;882:569-604.
7. Huang H, Wang C, Rubelt F, Scriba TJ, and Davis MM. Analyzing the Mycobacterium tuberculosis immune response by T-cell receptor clustering with GLIPH2 and genome-wide antigen screening. *Nat Biotechnol.* 2020;38(10):1194-202.
8. Hao Y, Hao S, Andersen-Nissen E, Mauck WM, 3rd, Zheng S, Butler A, et al. Integrated analysis of multimodal single-cell data. *Cell.* 2021;184(13):3573-87 e29.
9. Borcherding N, Bormann NL, and Kraus G. scRepertoire: An R-based toolkit for single-cell immune receptor analysis. *F1000Res.* 2020;9:47.
10. Huang Y, McCarthy DJ, and Stegle O. Vireo: Bayesian demultiplexing of pooled single-cell RNA-seq data without genotype reference. *Genome Biol.* 2019;20(1):273.
11. Pan M, and Li B. T cell receptor convergence is an indicator of antigen-specific T cell response in cancer immunotherapies. *Elife.* 2022;11.
12. Bergen V, Lange M, Peidli S, Wolf FA, and Theis FJ. Generalizing RNA velocity to transient cell states through dynamical modeling. *Nat Biotechnol.* 2020;38(12):1408-14.
13. La Manno G, Soldatov R, Zeisel A, Braun E, Hochgerner H, Petukhov V, et al. RNA velocity of single cells. *Nature.* 2018;560(7719):494-8.
14. Henderson LA, Volpi S, Frugoni F, Janssen E, Kim S, Sundel RP, et al. Next-Generation Sequencing Reveals Restriction and Clonotypic Expansion of Treg Cells in Juvenile Idiopathic Arthritis. *Arthritis Rheumatol.* 2016;68(7):1758-68.
15. Maschmeyer P, Heinz GA, Skopnik CM, Lutter L, Mazzoni A, Heinrich F, et al. Antigen-driven PD-1(+) TOX(+) BHLHE40(+) and PD-1(+) TOX(+) EOMES(+) T lymphocytes regulate juvenile idiopathic arthritis in situ. *Eur J Immunol.* 2021;51(4):915-29.
16. Sakuragi T, Yamada H, Haraguchi A, Kai K, Fukushi JI, Ikemura S, et al. Autoreactivity of Peripheral Helper T Cells in the Joints of Rheumatoid Arthritis. *J Immunol.* 2021;206(9):2045-51.
17. Fischer J, Dirks J, Klaussner J, Haase G, Holl-Wieden A, Hofmann C, et al. Effect of Clonally Expanded PD-1(high) CXCR5-CD4+ Peripheral T Helper Cells on B Cell Differentiation in the Joints of Patients With Antinuclear Antibody-Positive Juvenile Idiopathic Arthritis. *Arthritis Rheumatol.* 2022;74(1):150-62.
18. Musvosvi M, Huang H, Wang C, Xia Q, Rozot V, Krishnan A, et al. T cell receptor repertoires associated with control and disease progression following Mycobacterium tuberculosis infection. *Nat Med.* 2023;29(1):258-69.

19. Goncharov M, Bagaev D, Shcherbinin D, Zvyagin I, Bolotin D, Thomas PG, et al. VDJdb in the pandemic era: a compendium of T cell receptors specific for SARS-CoV-2. *Nat Methods.* 2022;19(9):1017-9.
20. Steere AC, Klitz W, Drouin EE, Falk BA, Kwok WW, Nepom GT, et al. Antibiotic-refractory Lyme arthritis is associated with HLA-DR molecules that bind a Borrelia burgdorferi peptide. *J Exp Med.* 2006;203(4):961-71.
21. Gonzalez-Galarza FF, McCabe A, Santos E, Jones J, Takeshita L, Ortega-Rivera ND, et al. Allele frequency net database (AFND) 2020 update: gold-standard data classification, open access genotype data and new query tools. *Nucleic Acids Res.* 2020;48(D1):D783-D8.
22. Klitz W, Maiers M, Spellman S, Baxter-Lowe LA, Schmeckpeper B, Williams TM, et al. New HLA haplotype frequency reference standards: high-resolution and large sample typing of HLA DR-DQ haplotypes in a sample of European Americans. *Tissue Antigens.* 2003;62(4):296-307.
23. Ausubel LJ, O'Connor KC, Baecher-Allen C, Trollmo C, Kessler B, Hekking B, et al. Characterization of in vivo expanded OspA-specific human T-cell clones. *Clin Immunol.* 2005;115(3):313-22.
24. Bondinas GP, Moustakas AK, and Papadopoulos GK. The spectrum of HLA-DQ and HLA-DR alleles, 2006: a listing correlating sequence and structure with function. *Immunogenetics.* 2007;59(7):539-53.
